# Next-Generation Sequencing Methods for Sensitive Hepatitis B Viral Genome Analysis: A European Study

**DOI:** 10.1101/2025.06.17.25329745

**Authors:** Michael X. Fu, Maria F. Perdomo, Sheila F. Lumley, Johan Ringlander, Kai Kean, Kaitlin Reid, Richard Mayne, Oscar Enrique Torres Montaguth, Leysa Forrest, Sarah Buddle, Johannes C. Botha, Joakim B. Stenbäck, Zachery Dickson, Chris Kent, Haiting Chai, Matthew Byott, Leo Hannolainen, Shannah Secret, George Airey, Klaus Hedman, Monique I. Andersson, M. Azim Ansari, Eleni Nastouli, Judith Breuer, Philippa C. Matthews, Tanya Golubchik, William L. Irving, Peter Simmonds, Heli Harvala

## Abstract

**Objectives:** This multicentre study investigated the utility of next-generation sequencing (NGS) to detect and generate hepatitis B virus (HBV) genomes in samples of low viral load (from 0.2 to 6207 IU/ml).

**Methods:** 23 HBV DNA positive plasma samples of genotypes A-E and one HBV-negative control sample were assayed blindly via 9 established NGS methods from 6 European laboratories. Methods included untargeted metagenomics, pre-enrichment by probe-capture followed by Illumina sequencing, and HBV-specific PCR pre-amplification followed by sequencing with Nanopore or Illumina.

**Results:** Full HBV genomes were obtained only from samples with viral loads >1000 IU/ml using probe-capture methods, >200 IU/ml using PCR-Illumina methods, >10 IU/ml using PCR-Nanopore methods, and in no samples using metagenomic methods. Contamination was observed in the negative control and samples with very low viral loads in all PCR-based methods. Probe-capture and metagenomic methods detected additional viruses not routinely screened in blood donations, including polyomaviruses and herpesviruses; positive results were confirmed by PCR.

**Conclusions:** NGS may delineate whole-genome sequences at low viral loads if supported by a PCR pre-amplification step. Probe-capture methods also reliably detect HBV without pre-amplification but achieve limited genome characterisation at low viral loads; they may additionally detect a wide range of blood-borne viruses.

## Introduction

The advent of high-throughput sequencing platforms in the mid-2000s, coined ‘next-generation sequencing’ (NGS), led to dramatic advances in the characterisation of host and pathogen genomes in health and disease^1^. Metagenomics, a commonly used NGS method to sequence all genomic material in a sample, has been instrumental in detecting broad-spectrum microbes, surveillance of emerging infections, and identifying novel pathogens^2,3^. Compared to Sanger sequencing where sub-genomic amplicons spanning regions of interest are sequenced, the ability to sequence complete microbial genomes using NGS in an unbiased manner has proven to be a powerful diagnostic tool^2,3^. However, the wider adoption of NGS in clinical microbiology and public health has been hindered by complex methodological workflows, complications in the detection and reporting of contaminating sequences, and a reduced sensitivity compared to real-time and nested PCR assays^2,4^. In particular, the inadequate sensitivity of untargeted metagenomics has limited the detection of hepatitis B virus (HBV), where many HBV-infected clinical samples contain low copy numbers of the viral genome^5,6^.

HBV remains a global public health issue despite the availability of a prophylactic vaccine and antiviral treatments; the virus chronically infects around 246 million individuals worldwide, accounting for 1.1 million HBV-related deaths in 2022^7^. In the blood, HBV exists in relaxed-circular DNA (rcDNA) form: a partially double-stranded genome of approximately 3.2 kb, consisting of overlapping reading frames encoding four genes: polymerase, surface, precore/core, and X. HBV can be classified into 10 genotypes (A to J) based upon sequence divergence ≥8%, with inter-genotypic differences in geographic distribution, transmission mode, and clinical outcomes^8^. Sequencing full-length HBV genomes identifies (sub)genotypes and recombinants and provides a better understanding of mechanisms of transmission and persistence, identifies treatment-resistant and immune-escape mutations, and thus can aid in the development of new therapeutics^9^. Furthermore, whole-genome sequencing (WGS) of low viral load (VL) samples enables the characterisation of occult HBV infection and the assessment of viral clearance, persistence and reactivation.

Methodologies used in clinical diagnostics for sequencing viral genomes need to be time-and cost-efficient, specific, and sensitive to support their integration into clinical, public health, and research laboratories^10^. Previous evaluations of viral sequencing methods have been limited to individual workflows and laboratories, with few coordinated comparisons, typically using synthetic or low-diversity materials due to limited clinical sample availability^11^. Method comparisons using clinical samples of varying loads are limited, but include a previous investigation of the effectiveness of NGS methods for the characterisation of hepatitis C virus detection frequencies and genome coverage^12^. The value of many studies is limited by the inclusion of samples with relatively high VLs without determining the detection limits of different NGS methods, for example^11^.

Amid calls for the HBV field to work progressively toward developing NGS methods^8^, this study brought together six European laboratories with nine established virus sequencing protocols, encompassing four commonly used NGS methods. First, the untargeted metagenomics method sequenced all genomic material in the samples. The second method involved the addition of biotinylated nucleic acid probes to enrich for large numbers of known pathogens and selectively capture target sequences before Illumina sequencing. The final methods involved PCR amplification of HBV fragments, followed by sequencing using the short-read Illumina or the long-read Oxford Nanopore Sequencing (ONT) platforms. All methods except untargeted metagenomics have previously demonstrated the ability to read and assemble whole HBV genomes from clinical samples^3,6,10,13^.

The four NGS methods were evaluated using a panel of 23 HBV DNA-positive plasma samples with low VLs. Through this comparison, we assessed the ability of NGS to detect and generate HBV sequences, as well as to co-detect other viruses potentially present in the samples. Overall, this study aims to provide an overview of the HBV-specific sensitivities of currently applied NGS methodologies and to evaluate their potential suitability for sequencing low VL samples in clinical practice.

## Materials and Methods

### Samples

Twenty-three large-volume samples, containing up to around 200 mL of plasma, from individuals infected with HBV genotypes A, B, C, D, and E were obtained from blood donors from NHS Blood and Transplant (NHSBT). To reflect the heterogeneity in the extraction methods used by NGS protocols and account for the non-negligible stochastic effect of measuring low VLs, geometric mean VLs were calculated across eight separate real-time PCR measurements from eight extractions (five of which were obtained from a previous investigation^14^ and three further extractions from 5 ml of plasma). The samples had geometric mean VLs ranging from 0.2 to 6207 IU/ml (approximately 1 to 31,000 copies/ml). Furthermore, a control plasma sample (around 200 mL) that screened negative for HBV serological markers, HBV DNA, and other blood-borne infections was obtained from NHSBT, where HBV DNA negativity was independently confirmed by real-time PCR measurements. All donors provided signed consent to use their samples for research purposes. This study was approved by the Blood Supply Clinical Audit, Risk and Effectiveness Committee of NHSBT (numbers 22031 and 23002). All 24 samples, including the negative control, were sent to each laboratory on dry ice, with anonymised sample identifiers and no other sample information.

### Sequencing methods

The panel of 24 blinded samples was used to evaluate the performance of nine established NGS protocols at six expert European laboratories, performed in the years 2023-2024. Methods included two untargeted metagenomic protocols using Illumina (MTG-A and MTG-B), three pan-viral probe-based enrichment protocols using Illumina (TAC-A, TAC-B, and TAC-C), two HBV-specific PCR amplification protocols using Illumina (PCR-A and PCR-B [PCR-Illumina]) and two HBV-specific PCR amplification protocols using ONT (PCR-1 and PCR-2 [PCR-ONT]). The protocols are specific to each laboratory, and the impact of individual methodological variables could not be evaluated independently within this study. Further details of protocols are found in the supplementary material. Of note, the input plasma volumes for extractions ranged from 0.2 to 20 ml. Extracted nucleic acids underwent library preparation according to local protocols, where input volumes ranged from 5 to 50 μl. Effective test volumes ranged from 4 to 800 μl. Average bases sequenced per sample differed from 2.2 Mb to 5.03 Gb per sample. The pre-capture libraries for TAC-B were sequenced for MTG-B; thus, the samples were prepared similarly for both protocols.

Factors relating to the technical practicality and costs associated with the use of NGS assays greatly influence their adoption for clinical diagnostics. Laboratories were asked to provide the total time and cost of each protocol step from extraction to sequencing on a standardised template document, considering the typical number of samples that would normally be included per run for each protocol. Times for each stage were separated into per-person labour time and waiting times (which included machine running times), and costs were standardised using a 1:0.90:0.75 USD:EUR:GBP exchange rate, as of May 18, 2025.

### Bioinformatic processing

Post-read filtering and genome assembly, either mapping to the closest available reference or *de novo*, were performed using established protocol-specific pipelines at each centre (see supplementary material). After deduplication of reads or removal of PCR duplicates and processing of sequences to begin at the EcoR1 restriction site (the conventional origin of the genome^15^), CSV files for each assembled genome/sample were sent from each centre, containing the number of base calls at each nucleotide site. These files were received at the coordinating laboratory, where further bioinformatics analyses were performed using a common set of tools on Microsoft Excel (version 16.103) and SSE^16,17^ (version 1.4) as previously described^12^. A 95% majority base consensus sequence was calculated at each genome position possessing one or more base reads, incorporating the calling of ambiguous bases when necessary. Constructed assembled sequences for each sample were examined and realigned to ensure consistent numbering, compare nucleotide similarities, and assess genome coverage and accuracy. Within-sample consensus sequences were assembled for each sample from the different NGS protocols by combining more than one similar assembled sequence with genome coverage of around 100% using the program Sequence Join in the SSE package^17^, followed by visual inspection of sites with ambiguous bases. Specific sequences from each method were compared with the within-sample consensus, and the numbers of nucleotide differences were recorded using the program Sequence Dist^17^. Missing bases were not included in comparisons of sequence identity. Thresholds for base-calls were set to practically represent the realities of sequencing: a threshold of one base read per nucleotide site was utilised for MTG and TAC methods, whilst a more stringent minimum threshold of 10 base reads at each nucleotide site was applied for PCR-based methods.

Within-sample consensus NGS sequences were (sub)genotyped as previously described^14^. When whole-genome assemblies could not be constructed (i.e. no two complete NGS sequences were similar), genotypes were defined from either the most complete NGS sequence or Sanger sequence, the latter if there were no similar NGS sequences. Individual NGS sequences from each protocol were genotyped using the Genome Detective system^18^.

### Nested PCR and Sanger sequencing

Sequences of a portion of the surface gene obtained previously from extensive nested PCR and Sanger sequencing were included for comparison to the NGS sequences^19^ (see Figure S1 for phylogenetic tree of all HBV-positive samples obtained via Sanger sequencing of nested PCR amplicons). Using DNA extracted from 5 ml of plasma^14^, nested PCR and sequencing of two further regions were performed: one spanning the surface and polymerase genes, and another spanning the X and precore/core genes (see Table S2 for primer details and nucleotide positions).

### Definitions

The following terms used in this manuscript have been defined to aid readability.

Protocol: unique individual NGS workflows performed by individual laboratories.

Method: a collection of similar protocols based on sequencing technology and platform. Methods included untargeted metagenomics followed by Illumina sequencing (MTG), targeted metagenomics using probe capture followed by Illumina sequencing (TAC), PCR amplification of HBV before Illumina sequencing (PCR-Illumina), and PCR amplification of HBV before ONT sequencing (PCR-ONT).

Genotype: distinct genetic variations of HBV based on nucleotide differences of ≥8%^8^.

Subgenotype: subgroups within specific HBV genotypes differing by 4-8% in viral genome sequence^8^ and distinct from the broader HBV genotypes.

Strain: a sequence that differed by >5% from other sequences from the same sample, indicating a different (sub)genotype. A 5% difference was chosen as a practical, rounded cutoff to reliably distinguish distinct subgenotypes.

Reference sequence: a known digital HBV DNA sequence used as a standard for genome assembly.

Assembled sequence: individual sequences constructed from individual protocols, which underwent read filtering and assembly via protocol-specific bioinformatic pipelines, followed by calculation of majority base consensus sequence at the coordinating laboratory and realignment of assembled sequences where necessary.

Within-sample consensus sequence: a more robust complete genome sequence for each sample, derived from the most commonly observed nucleotide at each genome position across available assembled sequences from all protocols for the same sample.

Contamination/contaminating sequence: a genetically different sequence and strain from the within-sample consensus sequence or other assembled sequences, where applicable (>5% nucleotide differences).

True-positive: similar to other sequences from the same sample (<5% nucleotide differences).

False-positive sequence: detection of an HBV DNA sequence in the negative control.

HBV-specific bases read/base counts: the total number of nucleotide bases aligned to the HBV genome, following deduplication of sequencing reads and removal of PCR duplicates where applicable.

Genome coverage: the proportion of the viral genome with consensus bases called.

Shannon entropy: a measure of the diversity of base counts in a sequence, which may result from technical sequencing errors or natural variability within infecting viral populations and could also be influenced by VL. Shannon entropy value ranges from 0 (no variability) to 1 (equal frequencies of all bases).

### Detection of other viruses by PCR

Since HBV was not the only possible virus sequenced from the five TAC and MTG protocols from three laboratories, the deduplicated read counts and genome coverage percentages were obtained for any additional viruses detected from the sample panel, following the full bioinformatics pipelines for each centre (see supplementary material for details of full pipelines). When a certain virus was detected by more than one NGS method in the same sample, confirmatory PCR was performed in all samples (see supplementary material for details of the PCR protocols).

### Statistical analyses

Spearman’s correlation was used to assess associations between VL and sequencing metrics across nine protocols. *P*-values were adjusted for multiple testing within each set of comparisons using the Benjamini-Hochberg method. α was 0.05. Data analyses and visualisation were performed with GraphPad Prism (v10.4.2, LLC), except *p*-value adjustment performed with Microsoft Excel (version 16.103).

## Results

### HBV detection and accuracy of assembled sequences

HBV amplicons were obtained for all 23 samples by Sanger sequencing, with good concordance between sequences generated by Sanger and NGS sequencing. Within-sample consensus NGS sequences were constructed for 14/15 samples with VL above 50 IU/ml (Table 1). Sequences assembled using different NGS protocols were generally identical or nearly identical to the within-sample consensus. A few individual sequences were >5% divergent from the within-sample consensus (shown in pink), indicating a different contaminating HBV strain. Assembled sequences were compared for eight samples with lower VLs, for which within-sample consensus sequences were unavailable. Different HBV genotypes were detected by at least one protocol for all eight samples (Table S3). Three samples, VLs 1.1, 1.8, and 8.5 IU/ml, yielded discrepant sequences in all NGS protocols.

**Table 1.**
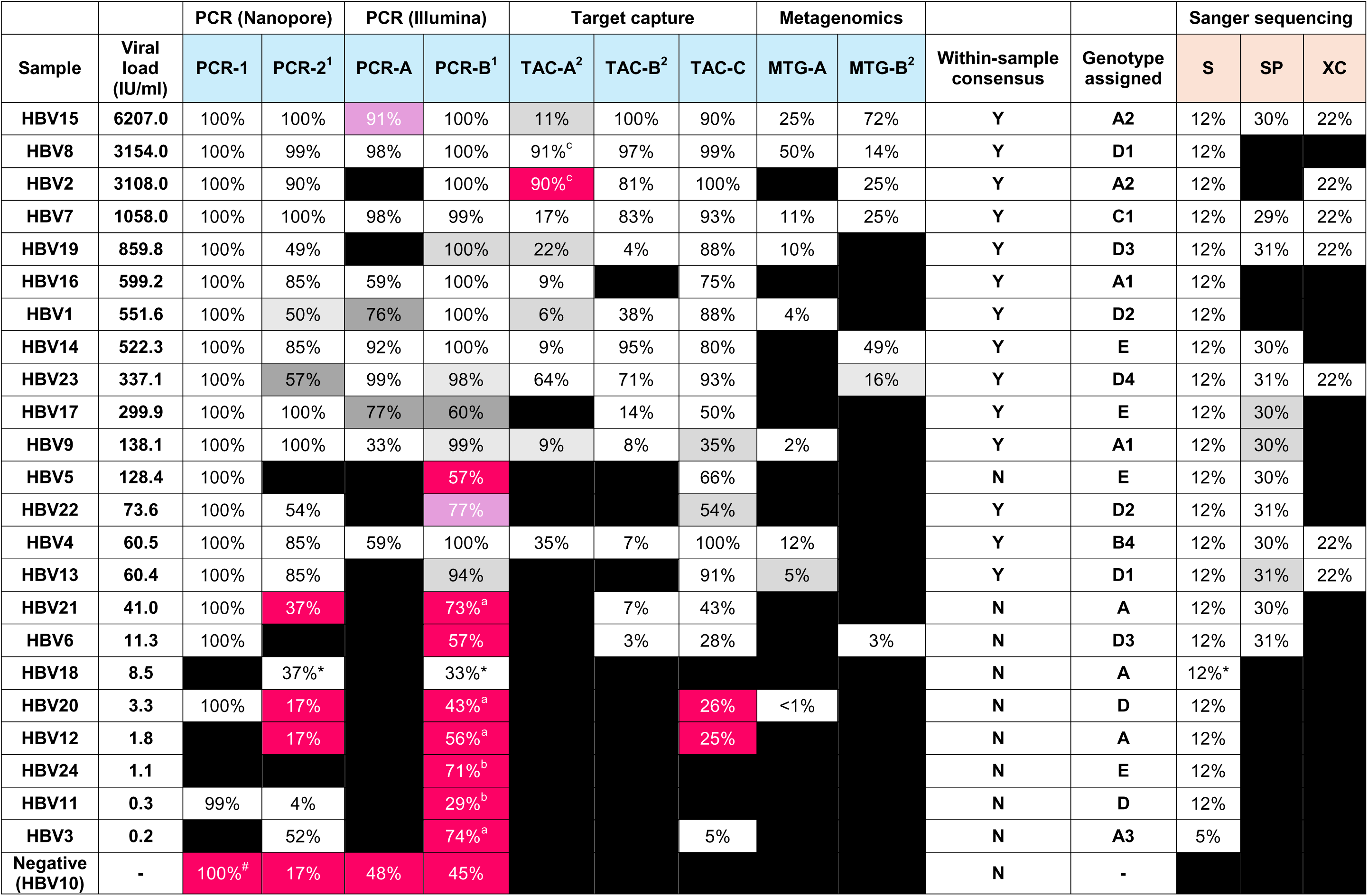
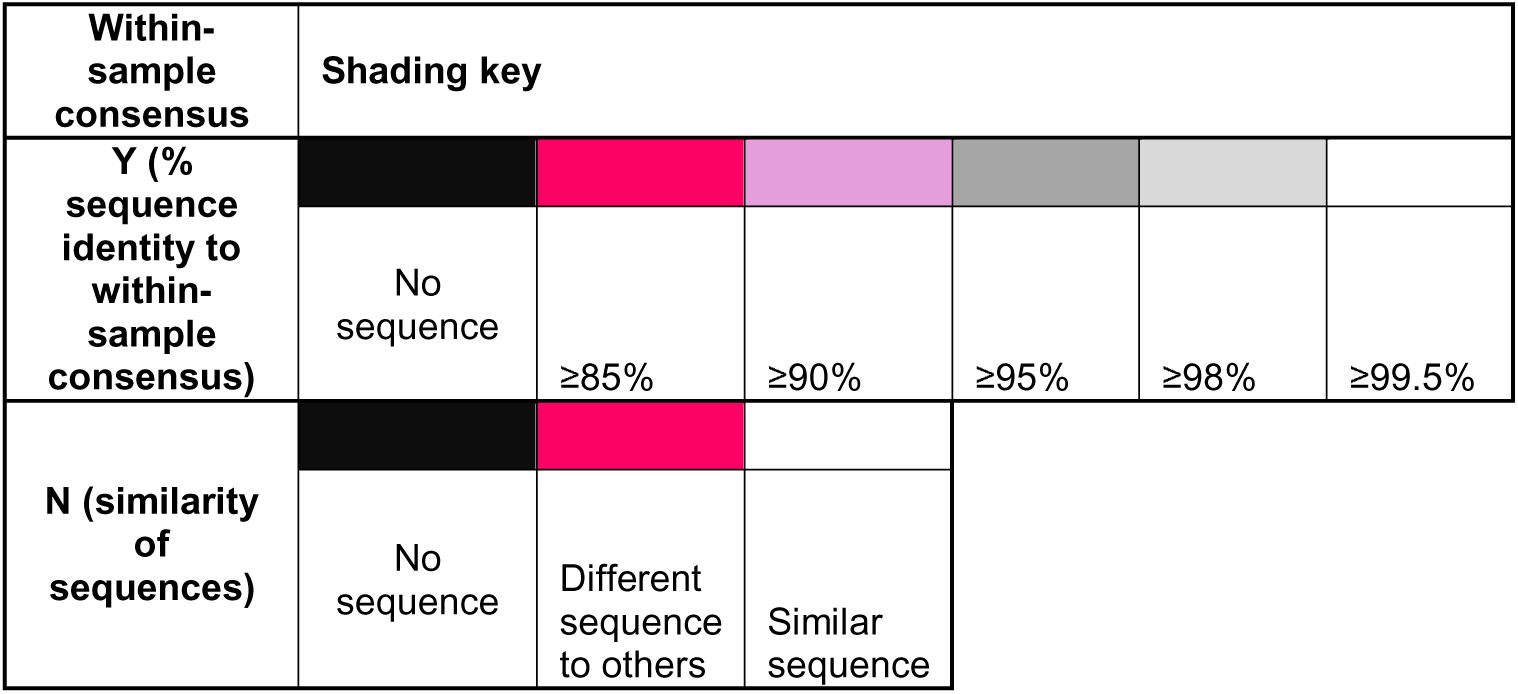
Overview of the detection of HBV in sample sets. The percentage genome completeness is given in each cell. The left-hand side of the table with blue headers shows the sequences obtained from the NGS methods, whilst the right-hand side (calamine colour) shows the sequences obtained by Sanger sequencing (length of S amplicon = 395 nucleotides; SP = 973 nucleotides; XC = 698 nucleotides). Within-sample consensus sequences were assembled for each sample from the individual sequences of the NGS methods that had around 100% genome coverage and were genetically similar, which were assigned to HBV genotypes. The percentage genetic relatedness of the sequences from each method was compared to the within-sample consensus sequence, where assembled sequences similar to the within-sample consensus were shaded on a grayscale, and different strains (differing in >5% nucleotide sequence from the within-sample consensus) were shaded in a pink scale. The absence of sequences was shaded in black. Where within-sample consensus sequences could not be assembled (i.e. no two complete sequences were similar), genotypes were defined from Sanger sequences, and the NGS sequences were instead compared to one another. Samples are ranked from highest to lowest geometric mean viral load. For TAC-B: library preparation failed for sample HBV13; samples HBV4, HBV9, and HBV19 had reads of poor quality for the original pipeline to process, so the mean base quality was dropped below 1.0. #: For PCR-1, a negative water control was tested instead of the negative plasma control. Alphabetical superscripts (a, b, c) represent identical sequences. Numerical superscripts next to method identifiers indicate where the same laboratory performed the protocols.

All four PCR-based protocols detected false-positive HBV sequences in the negative control (genome coverage, 48%-100%), which were identified as different genotypes (Table S3). Three of the four protocols also detected contaminating sequences in other samples, including those with low VL (<50 IU/ml). These four protocols were performed in three different laboratories, except for extraction, which the coordinating centre conducted for three of the four protocols. Some contaminants and false positives matched sequences of other *ex vivo* viruses (Table 1), but most were non-identical. For PCR-based methods other than PCR-1, the proportion of contaminants or the lack of detection at the lowest VLs deterred the ability to ascertain true-positive detection confidently. TAC methods had very few contaminating sequences, and no contaminating sequences were assembled from MTG methods.

Overall, true-positive HBV sequences of >5% genome coverage were detected in 5-6/23 samples using MTG, 10-17/23 samples using TAC, 9-13/23 samples using PCR-Illumina, and 16-19/23 samples using PCR-ONT protocols. MTG-B detected 7/13 samples at lower coverage and read numbers, which were detected by TAC-B, including the six samples with the highest coverage detected by TAC-B; samples for both protocols were prepared in the same run before the capture step.

### HBV base counts and genome coverage

The highest number of HBV-specific bases was read in PCR methods (up to 10^9^), followed by TAC (up to 10^7^) and the lowest in MTG (up to 10^4^). A strong correlation was observed between base counts and VL using the PCR-ONT and TAC methods, whereas this correlation was moderate when using the PCR-Illumina and MTG methods (Figure 1). Base counts were similar between samples for the PCR-Illumina methods. For PCR-ONT, bases obtained for true-positive samples with VL <50 IU/ml ranged from 1×10^3^ to 5×10^7^. For TAC and MTG methods, base counts were much lower for samples with lower VL. The approximate threshold to reliably detect the sample panel with true-positive sequences was >50 IU/ml or >10^6^ bases for PCR-ONT methods (except no threshold for PCR-1), >50 IU/ml or >10^4^ bases for TAC methods, >100 IU/ml or >10^7^ bases for PCR-Illumina methods, and >3000 IU/ml or >10^3^ bases for MTG methods.

**Figure 1.**
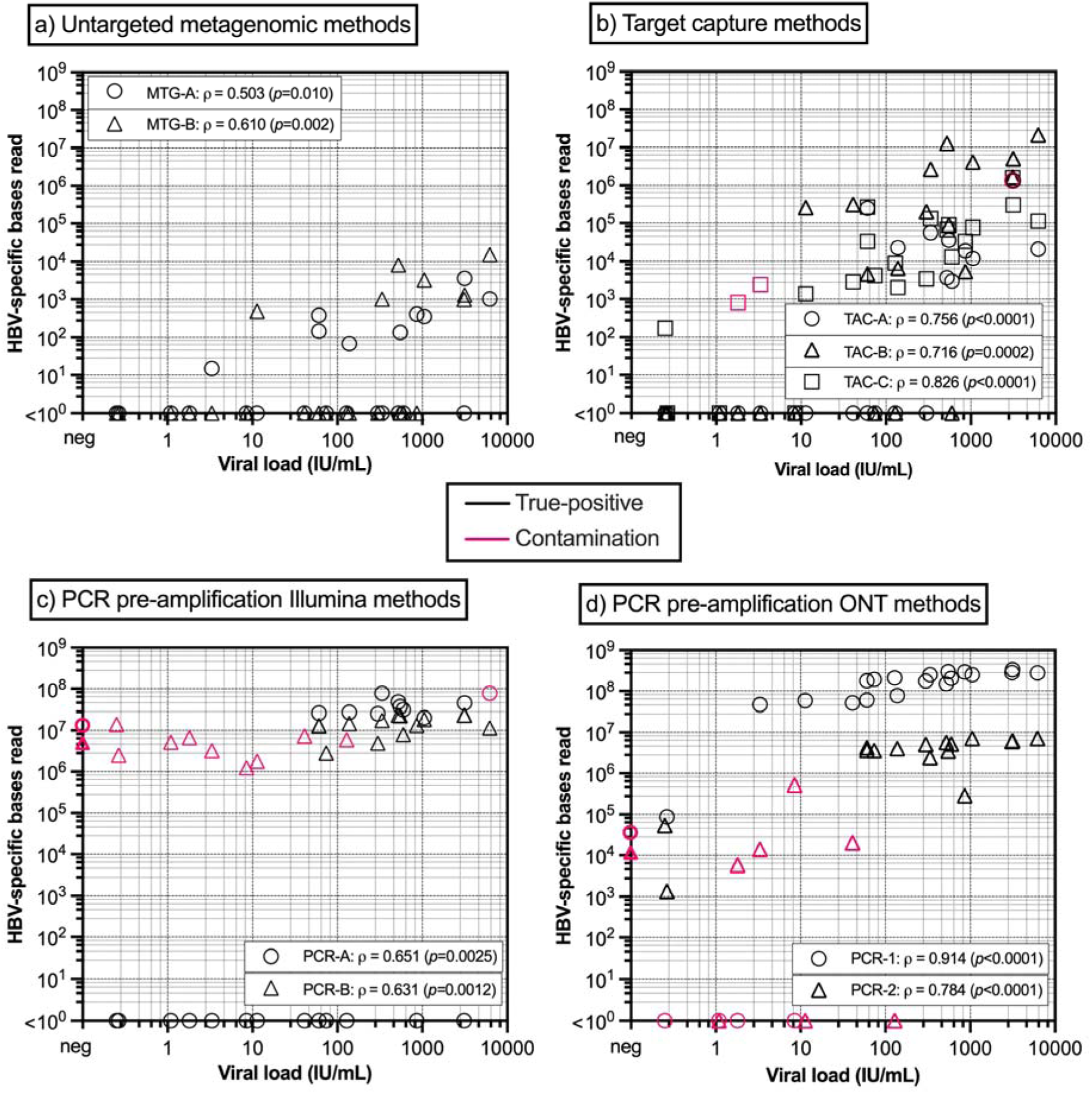
Relationship between viral loads and HBV-specific bases read for each group of methods on a common log scale. Pink indicates contaminating sequences, including where HBV sequences were obtained for the negative HBV10 sample on the y-axis. Samples not detected are found on the x-axis. Spearman’s correlation test with Benjamini-Hochberg adjustment deduced the significance of the association between linear viral loads and base counts (ρ value and *p*-values listed in boxes). HBV: hepatitis B virus; MTG: untargeted metagenomics; TAC: target capture; ONT: Oxford Nanopore Technologies.

Following multiple alignment of read assemblies, the completeness of assembled sequences was analysed. Complete or near-complete genome sequences were assembled for all samples with VL >10 IU/ml for PCR-1 protocol, >200 IU/ml for PCR-B, >1000 IU/ml for TAC-C, and in none of the samples for MTG methods (Figure 2). Coverage thresholds of 80% and 40% could be applied to exclude contamination in the PCR-Illumina and PCR-ONT methods, respectively, if the 100% coverage in the negative control from PCR-1 was excluded.

**Figure 2.**
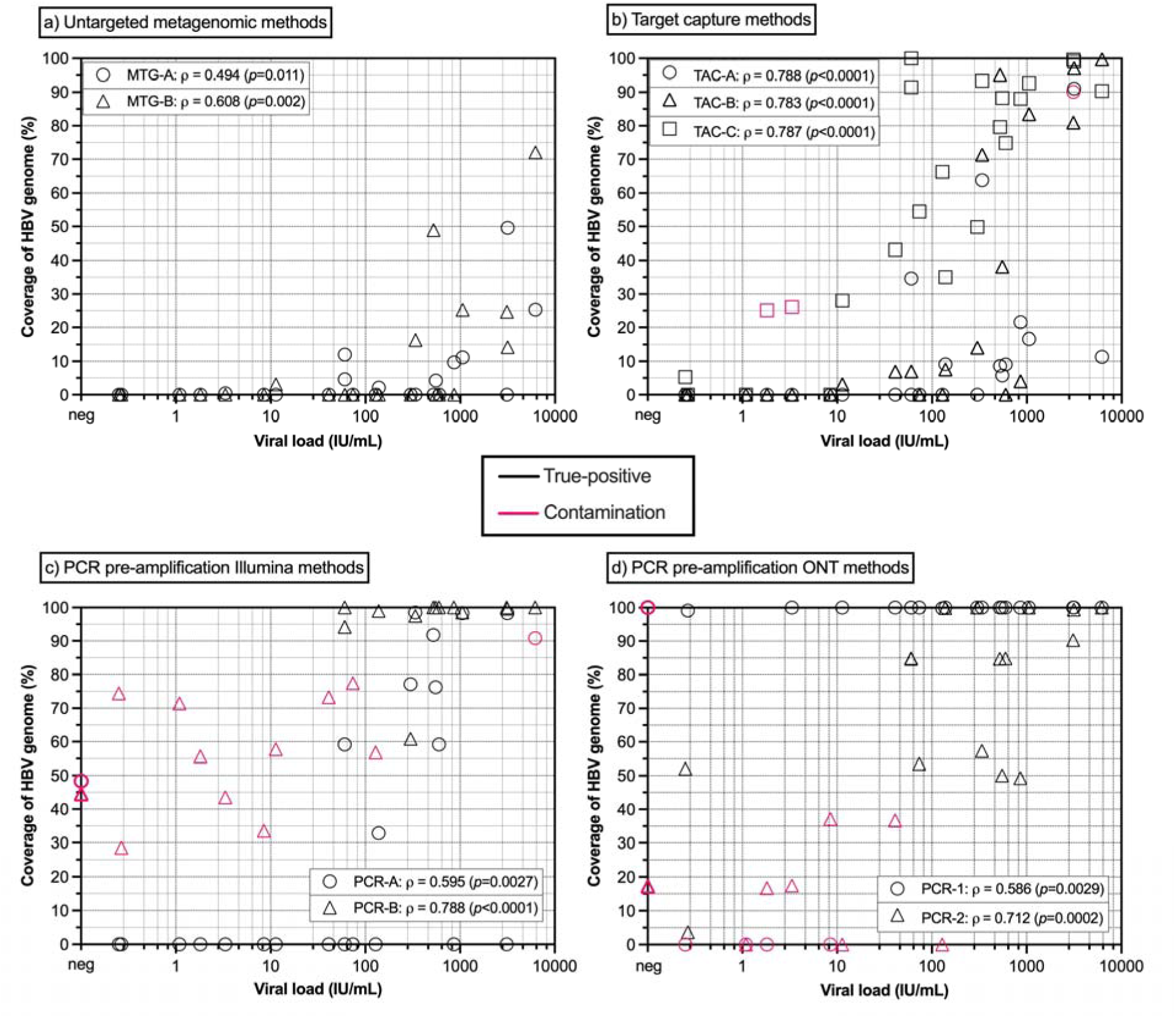
Relationship between viral loads and breadth coverage of HBV assembled sequences for each group of methods on a common axes scale. Pink indicates contaminating sequences, including where HBV sequences were obtained for the negative HBV10 sample on the y-axis. Samples not detected are found on the x-axis. Spearman’s correlation test with Benjamini-Hochberg adjustment deduced the significance of the association between linear viral loads and genome coverage (ρ value and *p*-values listed in boxes). HBV: hepatitis B virus; MTG: untargeted metagenomics; TAC: target capture; ONT: Oxford Nanopore Technologies.

Inspection of the mean genome coverage revealed that the methods yielded uneven coverage across the genome (Figure 3). PCR-ONT methods produced the most uniform genome coverage, with the exception of PCR-2, which showed reduced amplification spanning the region from the start of the polymerase to the surface genes. PCR-Illumina methods had more regions of reduced coverage, including genome positions 1100 to 2100 for PCR-A and the start of the surface gene for PCR-B. Probe capture efficiency was highly variable across protocols, each showing areas of markedly low capture efficiency. Coverage of the HBV genome was even sparser for MTG methods. When analysing individual samples, there was considerable heterogeneity in the distribution of HBV genome coverage between protocols, without clear patterns (Figure S2).

**Figure 3.**
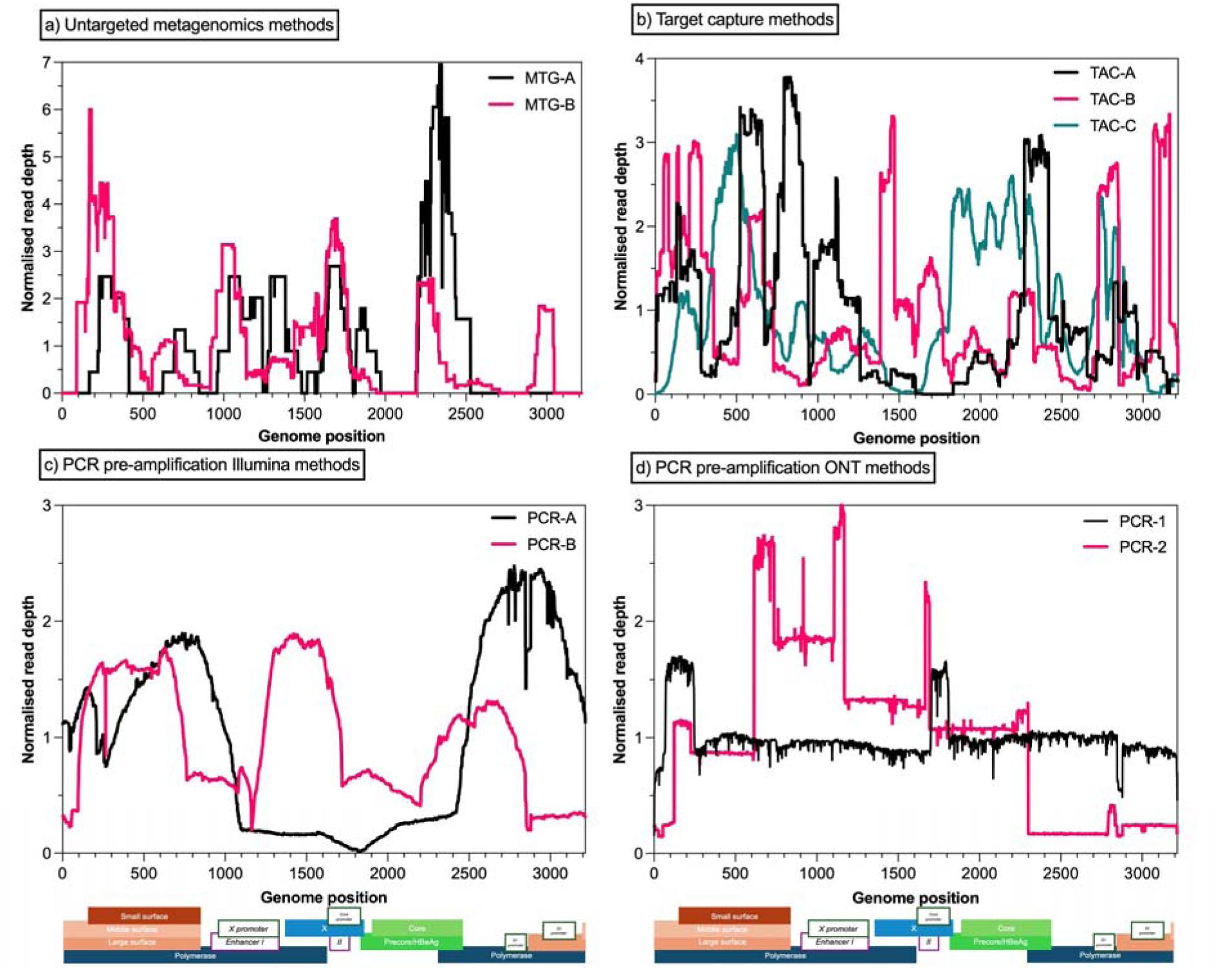
Normalised mean read depth of the HBV genome for each of the sequencing methods. The normalised mean read depth was calculated as the number of bases at each genome position as a proportion of the total reads of the sequence, then multiplied by the total number of sites, where the expected mean value was one base per site. The eleven highest viral load samples were included when detected, and only true-positive sequences with more than 1000 total HBV-specific bases read were included; the number of samples from each protocol that were included is indicated in each plot’s key. Genome positions were based on the D00330 reference sequence. A genome diagram of HBV, drawn to the x-axis scale, shows gene positions in shaded boxes and regulatory regions in unshaded boxes. TAC: target capture; MTG: untargeted metagenomics.

### Heterogeneity

The diversity of base counts at each site was quantified through calculations of Shannon entropy. Variability in Shannon values was observed between the NGS methods for the same samples, which may reflect the variations in the quantities of base counts by each method. MTG methods showed the least intra-sample HBV sequence heterogeneity, with a maximum mean Shannon entropy of 0.01, followed by the TAC methods (up to 0.04) and the PCR-Illumina methods (up to 0.09). Heterogeneity was extremely high (up to 0.32) for PCR-ONT methods (Figure 4). However, the presence of ambiguous bases in sequences from PCR-ONT methods did not affect the accuracy of consensus sequences generated from the reads. Diversity was higher when VL was greater for PCR-Illumina methods, but was lower when VL was higher for PCR-ONT methods.

**Figure 4.**
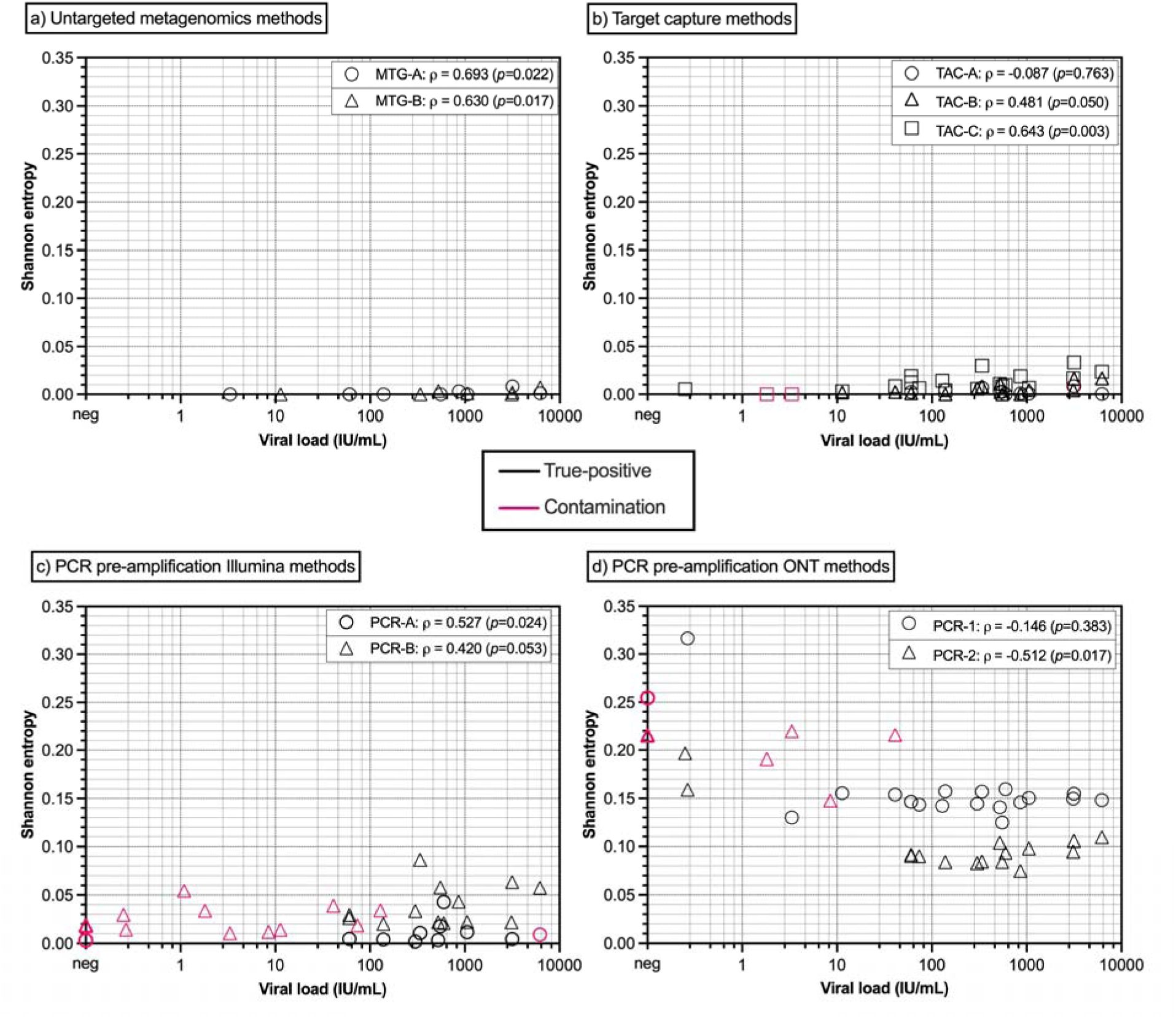
Relationship between viral loads and mean Shannon entropy values for polymorphic sites for each group of methods on a common axes scale. Pink indicates contaminating sequences, including where HBV sequences were obtained for the negative HBV10 sample on the y-axis. Samples not detected are found on the x-axis. Spearman’s correlation test with Benjamini-Hochberg adjustment deduced the significance of the association between linear viral loads and entropy values (ρ value and *p*-values listed in boxes). HBV: hepatitis B virus; MTG: untargeted metagenomics; TAC: target capture; ONT: Oxford Nanopore Technologies.

**Figure 5.**
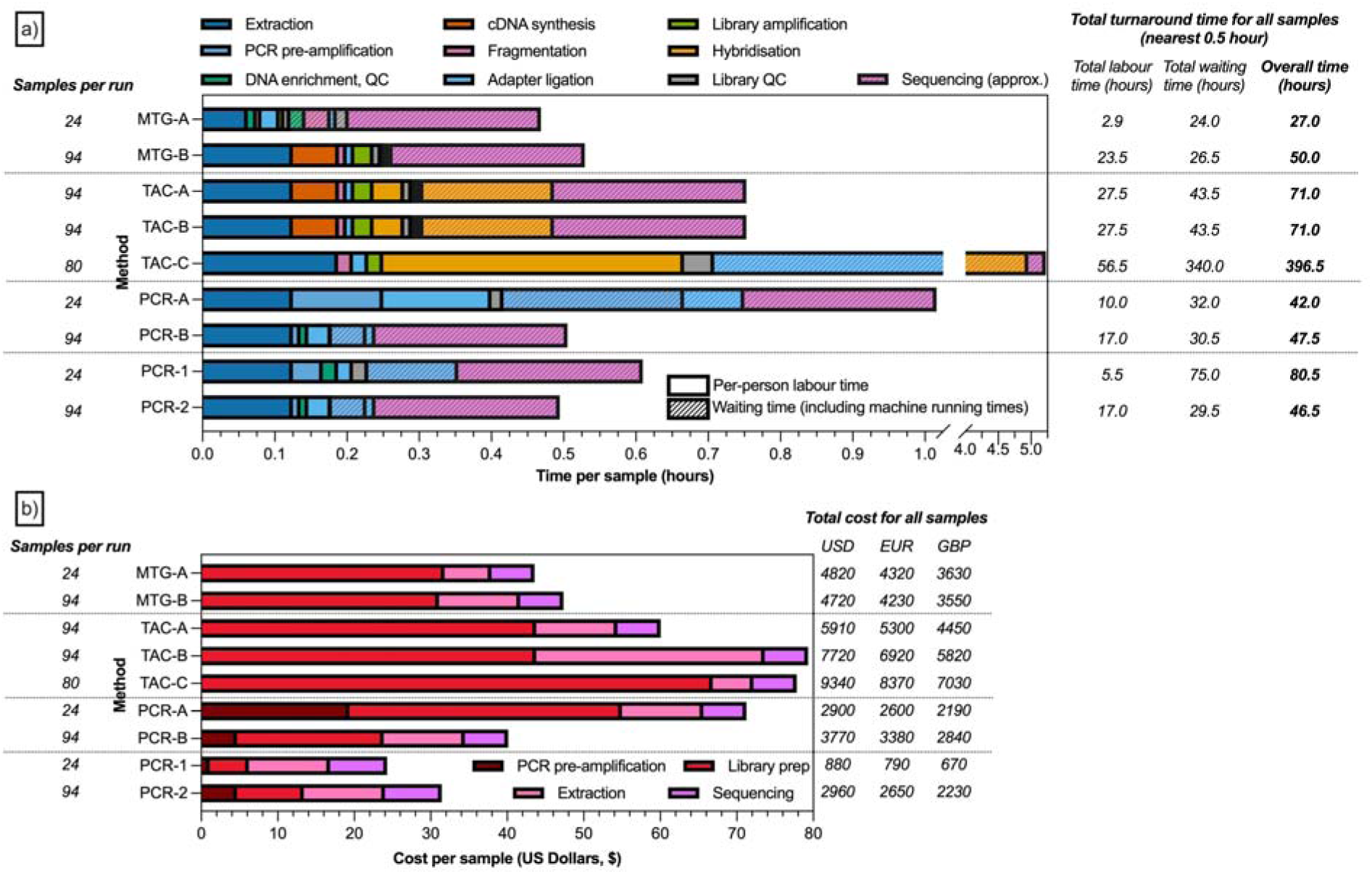
Comparison of a) time taken, and b) cost for each protocol based on the total time and costs for a protocol-specific representative number of samples included in a typical run. For Figure 5a, coloured bars to the left of the plot show per-person labour time per sample, whereas the same-coloured bars with lighter-coloured diagonal lines to the right show the waiting time per sample, which may include machine running times. For visual comparison, sequencing time and costs per sample were standardised to the shortest and cheapest Illumina or Nanopore protocol; the overall times reported include the full sequencing durations as recorded by each laboratory. For clinical purposes, PCR-1 is normally run overnight on the MinIon flow cell (average of 16 hours), but was run for 72 hours for this study. Costs were standardised using a 1:0.8959:0.7531 USD:EUR:GBP exchange rate, as of May 18, 2025, and total costs were rounded to the nearest ten. USD: United States Dollar; EUR: Euro; GBP: Great British Pound; QC: quality control; TAC: target capture; MTG: untargeted metagenomics.

### Co-detection

Protocol-specific bioinformatic pipelines were employed to determine the presence, read counts, and genome coverage of other viruses for the TAC and MTG methods, which simultaneously screened for multiple viruses from generated sequences (Table 2). Altogether, anellovirus sequences were detected in 22/24 samples by the five NGS protocols. Anellovirus was detected by at least two protocols in 19/24 samples, and 2/24 samples by all five protocols. >50% genome coverage for anellovirus was observed in two samples although consensus sequence generation was hampered by the likely presence of multiple strains and types of anelloviruses within samples. Confirmatory PCR for alpha, beta, and gammatorqueviruses revealed the presence of DNA sequences of all three anellovirus genera in all 24 samples (20/24 αTTV-positive αTTV; 22/24 βTTV-positive; 17/24 γTTV-positive), all with relatively high VLs (Ct values ranged from 4.7 to 26.1). Sanger sequencing of amplicons generated by nested PCR from randomly selected anellovirus-positive samples confirmed the presence of different strains from each genus.

**Table 2.**
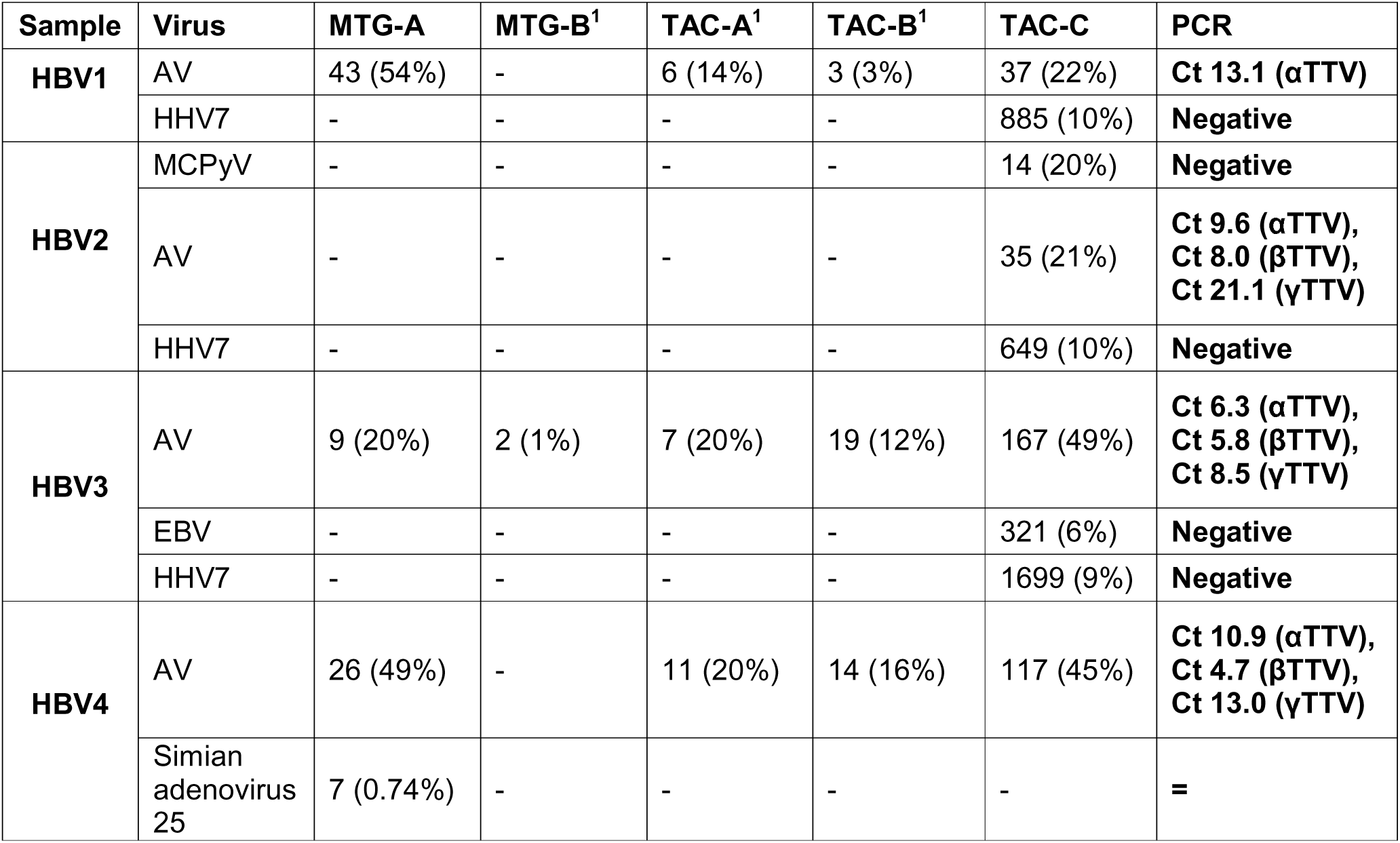
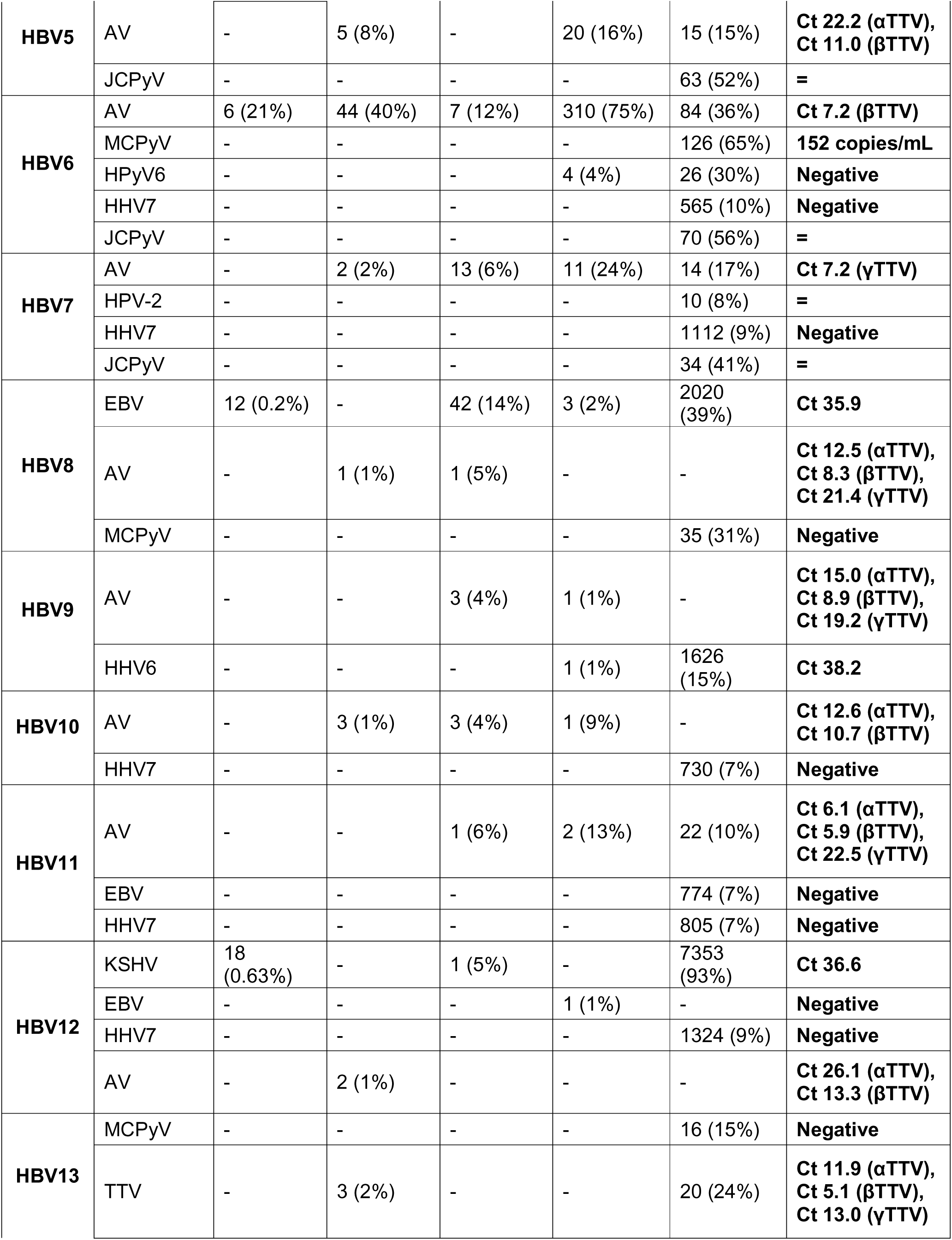
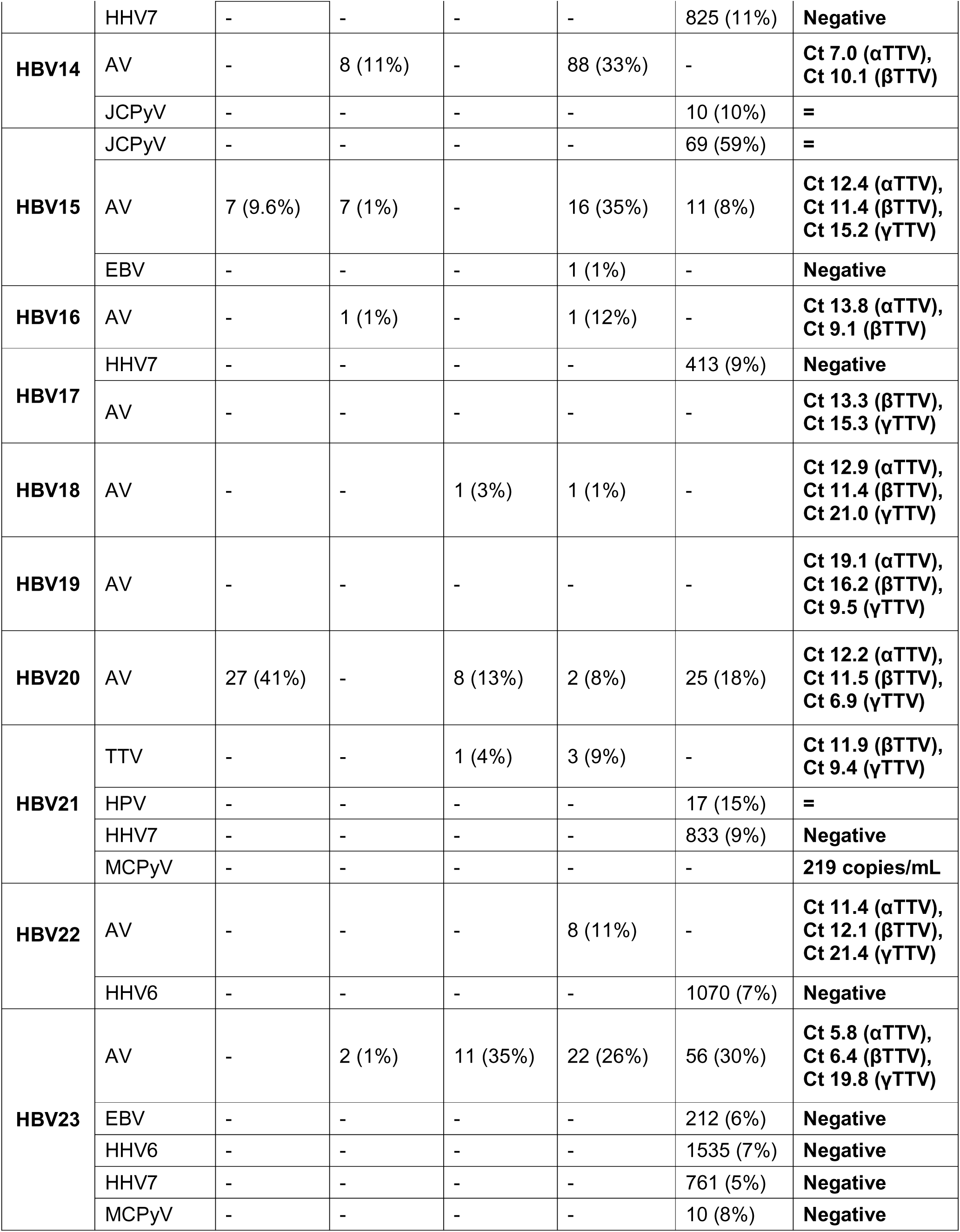
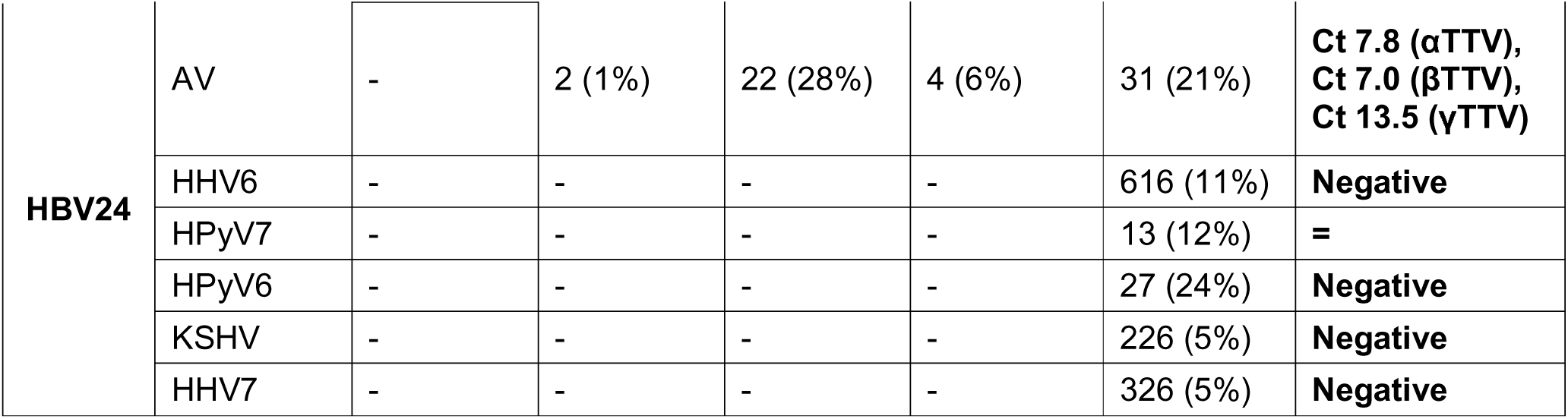
Detection of other viruses by the targeted capture and untargeted metagenomics methods, showing deduplicated read counts (genome coverage %). Results reported for TAC-C are for viruses where at least 10 reads were obtained. More than one TTV species may have been identified for MTG-A, but the most abundant was included. The furthest right column shows the PCR results, where PCR was performed for all samples when two or more methods detected a virus in the same sample. These PCRs were specific for alpha, beta and gamma-torquevirus, Merkel cell polyomavirus, and herpesviruses. ‘-’ denotes where the method did not detect the virus, ‘=’ denotes where the PCR was not performed. A numerical superscript indicates where the same laboratory performed the protocols. HHV: human herpesvirus; MCPyV: Merkel cell polyomavirus; AV: anellovirus; TTV: torquetenovirus; EBV: Epstein-Barr virus; KSHV: Kaposi’s sarcoma-associated herpesvirus; JCPyV: John Cunningham virus.

| Sample | Virus | MTG-A | MTG-B<sup>1</sup> | TAC-A<sup>1</sup> | TAC-B<sup>1</sup> | TAC-C | PCR |
| --- | --- | --- | --- | --- | --- | --- | --- |
| **HBV1** | AV | 43 (54%) | - | 6 (14%) | 3 (3%) | 37 (22%) | **Ct 13.1 (αTTV)** |
|  | HHV7 | - | - | - | - | 885 (10%) | **Negative** |
| **HBV2** | MCPyV | - | - | - | - | 14 (20%) | **Negative** |
|  | AV | - | - | - | - | 35 (21%) | **Ct 9.6 (αTTV),**<br>**Ct 8.0 (βTTV),**<br>**Ct 21.1 (γTTV)** |
|  | HHV7 | - | - | - | - | 649 (10%) | **Negative** |
| **HBV3** | AV | 9 (20%) | 2 (1%) | 7 (20%) | 19 (12%) | 167 (49%) | **Ct 6.3 (αTTV),**<br>**Ct 5.8 (βTTV),**<br>**Ct 8.5 (γTTV)** |
|  | EBV | - | - | - | - | 321 (6%) | **Negative** |
|  | HHV7 | - | - | - | - | 1699 (9%) | **Negative** |
| **HBV4** | AV | 26 (49%) | - | 11 (20%) | 14 (16%) | 117 (45%) | **Ct 10.9 (αTTV),**<br>**Ct 4.7 (βTTV),**<br>**Ct 13.0 (γTTV)** |
|  | Simian<br>adenovirus<br>25 | 7 (0.74%) | - | - | - | - | **=** |
| **HBV5** | AV | - | 5 (8%) | - | 20 (16%) | 15 (15%) | **Ct 22.2 (αTTV),**<br>**Ct 11.0 (βTTV)** |
|  | JCPyV | - | - | - | - | 63 (52%) | **=** |
| **HBV6** | AV | 6 (21%) | 44 (40%) | 7 (12%) | 310 (75%) | 84 (36%) | **Ct 7.2 (βTTV)** |
|  | MCPyV | - | - | - | - | 126 (65%) | **152 copies/mL** |
|  | HPyV6 | - | - | - | 4 (4%) | 26 (30%) | **Negative** |
|  | HHV7 | - | - | - | - | 565 (10%) | **Negative** |
|  | JCPyV | - | - | - | - | 70 (56%) | **=** |
| **HBV7** | AV | - | 2 (2%) | 13 (6%) | 11 (24%) | 14 (17%) | **Ct 7.2 (γTTV)** |
|  | HPV-2 | - | - | - | - | 10 (8%) | **=** |
|  | HHV7 | - | - | - | - | 1112 (9%) | **Negative** |
|  | JCPyV | - | - | - | - | 34 (41%) | **=** |
| **HBV8** | EBV | 12 (0.2%) | - | 42 (14%) | 3 (2%) | 2020 (39%) | **Ct 35.9** |
|  | AV | - | 1 (1%) | 1 (5%) | - | - | **Ct 12.5 (αTTV),**<br>**Ct 8.3 (βTTV),**<br>**Ct 21.4 (γTTV)** |
|  | MCPyV | - | - | - | - | 35 (31%) | **Negative** |
| **HBV9** | AV | - | - | 3 (4%) | 1 (1%) | - | **Ct 15.0 (αTTV),**<br>**Ct 8.9 (βTTV),**<br>**Ct 19.2 (γTTV)** |
|  | HHV6 | - | - | - | 1 (1%) | 1626 (15%) | **Ct 38.2** |
| **HBV10** | AV | - | 3 (1%) | 3 (4%) | 1 (9%) | - | **Ct 12.6 (αTTV),**<br>**Ct 10.7 (βTTV)** |
|  | HHV7 | - | - | - | - | 730 (7%) | **Negative** |
| **HBV11** | AV | - | - | 1 (6%) | 2 (13%) | 22 (10%) | **Ct 6.1 (αTTV),**<br>**Ct 5.9 (βTTV),**<br>**Ct 22.5 (γTTV)** |
|  | EBV | - | - | - | - | 774 (7%) | **Negative** |
|  | HHV7 | - | - | - | - | 805 (7%) | **Negative** |
| **HBV12** | KSHV | 18 (0.63%) | - | 1 (5%) | - | 7353 (93%) | **Ct 36.6** |
|  | EBV | - | - | - | 1 (1%) | - | **Negative** |
|  | HHV7 | - | - | - | - | 1324 (9%) | **Negative** |
|  | AV | - | 2 (1%) | - | - | - | **Ct 26.1 (αTTV),**<br>**Ct 13.3 (βTTV)** |
| **HBV13** | MCPyV | - | - | - | - | 16 (15%) | **Negative** |
|  | TTV | - | 3 (2%) | - | - | 20 (24%) | **Ct 11.9 (αTTV),**<br>**Ct 5.1 (βTTV),**<br>**Ct 13.0 (γTTV)** |
|  | HHV7 | - | - | - | - | 825 (11%) | **Negative** |
| **HBV14** | AV | - | 8 (11%) | - | 88 (33%) | - | **Ct 7.0 (αTTV),**<br>**Ct 10.1 (βTTV)** |
|  | JCPyV | - | - | - | - | 10 (10%) | **=** |
| **HBV15** | JCPyV | - | - | - | - | 69 (59%) | **=** |
|  | AV | 7 (9.6%) | 7 (1%) | - | 16 (35%) | 11 (8%) | **Ct 12.4 (αTTV),**<br>**Ct 11.4 (βTTV),**<br>**Ct 15.2 (γTTV)** |
|  | EBV | - | - | - | 1 (1%) | - | **Negative** |
| **HBV16** | AV | - | 1 (1%) | - | 1 (12%) | - | **Ct 13.8 (αTTV),**<br>**Ct 9.1 (βTTV)** |
| **HBV17** | HHV7 | - | - | - | - | 413 (9%) | **Negative** |
|  | AV | - | - | - | - | - | **Ct 13.3 (βTTV),**<br>**Ct 15.3 (γTTV)** |
| **HBV18** | AV | - | - | 1 (3%) | 1 (1%) | - | **Ct 12.9 (αTTV),**<br>**Ct 11.4 (βTTV),**<br>**Ct 21.0 (γTTV)** |
| **HBV19** | AV | - | - | - | - | - | **Ct 19.1 (αTTV),**<br>**Ct 16.2 (βTTV),**<br>**Ct 9.5 (γTTV)** |
| **HBV20** | AV | 27 (41%) | - | 8 (13%) | 2 (8%) | 25 (18%) | **Ct 12.2 (αTTV),**<br>**Ct 11.5 (βTTV),**<br>**Ct 6.9 (γTTV)** |
| **HBV21** | TTV | - | - | 1 (4%) | 3 (9%) | - | **Ct 11.9 (βTTV),**<br>**Ct 9.4 (γTTV)** |
|  | HPV | - | - | - | - | 17 (15%) | **=** |
|  | HHV7 | - | - | - | - | 833 (9%) | **Negative** |
|  | MCPyV | - | - | - | - | - | **219 copies/mL** |
| **HBV22** | AV | - | - | - | 8 (11%) | - | **Ct 11.4 (αTTV),**<br>**Ct 12.1 (βTTV),**<br>**Ct 21.4 (γTTV)** |
|  | HHV6 | - | - | - | - | 1070 (7%) | **Negative** |
| **HBV23** | AV | - | 2 (1%) | 11 (35%) | 22 (26%) | 56 (30%) | **Ct 5.8 (αTTV),**<br>**Ct 6.4 (βTTV),**<br>**Ct 19.8 (γTTV)** |
|  | EBV | - | - | - | - | 212 (6%) | **Negative** |
|  | HHV6 | - | - | - | - | 1535 (7%) | **Negative** |
|  | HHV7 | - | - | - | - | 761 (5%) | **Negative** |
|  | MCPyV | - | - | - | - | 10 (8%) | **Negative** |
| **HBV24** | AV | - | 2 (1%) | 22 (28%) | 4 (6%) | 31 (21%) | **Ct 7.8 (αTTV),**<br>**Ct 7.0 (βTTV),**<br>**Ct 13.5 (γTTV)** |
|  | HHV6 | - | - | - | - | 616 (11%) | **Negative** |
|  | HPyV7 | - | - | - | - | 13 (12%) | **=** |
|  | HPyV6 | - | - | - | - | 27 (24%) | **Negative** |
|  | KSHV | - | - | - | - | 226 (5%) | **Negative** |
|  | HHV7 | - | - | - | - | 326 (5%) | **Negative** |

Various human herpesviruses were detected in 17/24 samples across the five MTG and TAC protocols at <50% genome coverage, and were detected by more than one protocol for three samples (EBV in 4/5 protocols in two laboratories for sample HBV8; HHV6 in 2/5 protocols in two laboratories for HBV9; KSHV in 3/5 protocols in three laboratories for HBV12). Real-time PCR confirmed the presence of these herpesviruses in these three samples, but no other sample was HHV PCR-positive, including the 13 samples where TAC-C reported HHV-7 DNA sequences. Polyomavirus DNA sequences were detected in 11/24 samples using NGS protocols; however, they were only detected by more than one protocol in one sample (HPyV6, detected by two protocols in two laboratories in HBV6). HyPV6 was, however, not detected by real-time PCR. Instead, MCPyV was detected by real-time PCR in low copy numbers in HBV6 (detected by one NGS protocol) and HBV21 (not detected by any NGS protocol). Overall, TAC methods detected a greater number of viruses at higher read counts and coverage than MTG methods.

### Time and costs

The per-person labour and waiting times varied between protocols (Figure 4a). The turnaround time was shortest for MTG and most PCR-based protocols. TAC methods took longer, mostly due to overnight hybridisation steps. Costs were lowest for PCR-ONT methods, higher for PCR-Illumina methods, followed by MTG methods, whilst TAC methods had the greatest cost (Figure 4b).

## Discussion

Findings from our multicentre study provide an overview of currently available NGS methodologies for sequencing low VL HBV clinical samples. They highlight the HBV-specific sensitivities and potential applicability in clinical practice. While all evaluated methods demonstrated varying capacities to generate HBV sequences and detect co-infecting viruses, protocols incorporating pre-amplification steps showed greater sensitivity at low VLs. Reliable detection of HBV at VL ∼10 IU/ml (for PCR-1 and TAC-C) or ∼100 IU/ml (for other pre-amplification protocols) would be sufficient for most HBV infections encountered in clinical settings. For detecting and quantifying samples with very low VL, such as those associated with occult HBV^14^, conventional real-time or nested PCR remained essential. Previous investigations of PCR-1 and PCR-2 also showed successful amplification of full HBV genomes at VL <30 IU/ml^6,13^, and alternative probe-capture methods observed a similar 50 copies/ml limit of detection for HBV^20^ and 90-100% coverage for a range of DNA and RNA viruses at ≥1000 copies/ml^21,22^. The substantially reduced depth of coverage for MTG methods we observed has been well-described for a range of viruses^10–12^. However, the method retains its value in other applications due to its ability to detect unanticipated or potentially entirely novel pathogens without a requirement to pre-select targets in capture and PCR-assisted NGS methods^2^.

Although detection sensitivity was similar, genomes generated from PCR-based methods were near-complete compared to lower genome coverage from TAC methods for our low VL samples. As an inexpensive method for sequencing whole HBV genomes, PCR-ONT NGS may be a beneficial screening tool for the HBV and microbiology fields, especially in low- and middle-income countries^23^ when ONT allows preliminary analysis before a run is completed: highly advantageous in terms of turnaround time for high VL samples. Our findings contrast with a similar study on hepatitis C virus, which concluded that PCR-based NGS was relatively laborious^12^; improvements in technologies and workflows could explain the differences found. However, PCR-based NGS would be more challenging for more divergent and/or larger viruses than HBV, as primer design for whole-genome amplification can be difficult, and amplicon drop-out necessitating primer re-design is not uncommon. Variables that may have affected the methods’ detection capabilities in this study include extraction from larger volumes of plasma (as previously found with PCR-1^13^ and TAC-A/B protocols^19^), having greater sample representation and fewer samples for library preparation, and sequencing more gigabases per sample. For PCR-based methods, sensitivity is affected less by the sequencing platform and more by the effectiveness of the PCR for genome pre-amplification, particularly the conservation of primer-binding sites. A nested PCR strategy with shorter amplicons would improve sensitivity if WGS is not required. For TAC methods, sensitivity may have been affected by factors such as probe design, hybridisation, and wash temperatures. As with the detection of viruses around the limit of detection of conventional PCR assays, the stochastic effect around the limits of detection of NGS protocols affects the detection of samples with low VL, albeit at a higher detection limit than conventional PCR.

Although the sensitivity of PCR-based NGS methods is sufficient for most clinical cases of HBV, the finding of low-level HBV DNA contamination compromised their diagnostic reliability at the lowest VLs. Without applying a minimum threshold of 10 for consensus base calling to account for potential background contamination or sequencing errors, HBV sequences would have been detected in all samples (24/24) using PCR-based methods; however, contaminant HBV sequences would also have appeared more frequently (Table S4). The detection of contaminating sequences has been reported in other studies comparing NGS protocols^11,21^, highlighting contamination as a persistent obstacle to the reliable characterisation of low VL genomes. Notably, one planned protocol could not be included in this study because the background level of contamination exceeded the VLs of the sample panel.

Investigating the exact sources and mechanisms of contamination was beyond the scope of this study. The negative control was confirmed to be HBV-negative prior to processing, suggesting that contamination likely occurred during library preparation, amplification, or sample handling in each of the participating laboratories. This interpretation is supported by the detection of identical sequences across multiple samples processed within the same protocol or laboratory (Table 1). The variation observed in both the number and characteristics of contaminant strains among protocols indicates that contamination events in low VL samples are relatively frequent but highly context-dependent. In the presence of background contamination, stricter analytical thresholds can reduce sensitivity but improve the reliability of positive calls. Such measures may include increasing the minimum read depth required for consensus base calls, enforcing minimum genome coverage or VL thresholds, and setting defined read count criteria relative to negative controls (Table S5). Importantly, minimising contamination in complex NGS workflows requires extensive experience and rigorous laboratory practice^24^, such as using separate PCR workspaces and tubes, exercising meticulous pipetting, and including appropriate negative plasma controls.

The substantial variation in base counts and error rates observed among ONT base-calling algorithms used between the years 2023 and 2024 made it difficult to accurately identify minority variants. Although these errors are unlikely to impact interpretation for most clinical and public health purposes, we advise against performing variant calling directly from mapped ONT reads without considerable bioinformatics expertise to exclude false substitutions and improve variant calling^25^. Subsequent improvements in R10 flow cells and base-calling algorithms, after our experiments were performed, are likely to enhance sequencing accuracy and reduce base count variability^26,27^.

NGS methods involving PCR amplification are inherently limited to the characterisation of one known target pathogen. Their use for whole genome characterisation is compromised for viruses with long genomes such as herpesviruses, where multiple, overlapping PCRs may be required, or for targets which are highly genetically heterogeneous and which may lack conserved sites for primer binding across the genome, such as anelloviruses. Metagenomic methods possess clear advantages in their breadth of detection, with frequent co-detections of multiple anelloviruses, human herpesviruses, and polyomaviruses in the samples. The 100% (24/24) prevalence of anellovirus DNA detection by PCR was consistent with previous studies, which have shown that most immunocompetent individuals carry the virus in their blood and at high VLs in patients with HBV infection^28^. The low detection rate by all TAC protocols may have originated from the extreme genetic heterogeneity of the anellovirus species and genera that were not adequately represented among the probes used for target capture^29^. However, this does not explain the similarly low rates of detection by MTG methods, which were not a sensitivity problem as the anelloviruses had high VLs by PCR; it is possible that their circular sequences were too divergent to map to what might be a very limited number of reference sequences used by the bioinformatic software to assign reads. Our detection rates of polyomaviruses and HHVs were comparable to those reported in population studies^30,31^. The high detection rate of HHVs and polyomaviruses with TAC-C compared to fragmented PCR could potentially be explained by the presence of incomplete or highly fragmented genome sequences derived from degraded viral DNA from disintegrating persistently infected cells^32^.

Our study focused on HBV samples with low VLs; including samples with higher VLs would likely have increased the proportion achieving high base counts and complete genome coverage, while reducing the relative impact of contamination on detection reliability. Although we centralised some of the bioinformatics to facilitate comparison, the different pipeline-specific steps performed by each laboratory could have affected sensitivity, such as the choice of taxonomic classifier^10^. In a follow-up study, we plan to evaluate the performance of different bioinformatic pipelines on the raw data generated in the current study to ascertain optimal tools for HBV characterisation.

In conclusion, this study provides an overview of the HBV-specific sensitivities of currently used NGS methodologies and their potential applicability for sequencing low VL clinical samples. NGS approaches incorporating PCR amplification demonstrate sufficient sensitivity for most HBV infections typically encountered in clinical practice, where VLs exceed approximately 10 IU/ml. However, the presence of low-level contamination remains a key limitation, restricting reliable genome identification at the lowest VLs. Future efforts should focus on identifying and mitigating sources of background contamination to enhance the specificity and reliability of NGS. Strengthening contamination control would enable broader clinical implementation of NGS as a valuable tool for characterising HBV and other viral pathogens in diverse diagnostic settings.

## Acknowledgements

The authors thank Dr Belinda Singleton (NHS Blood and Transplant) for helping develop the anellovirus PCR protocol. This study was primarily supported by the National Institute for Health and Care Research [grant number NIHR203338], which supported these authors under the Blood and Transplant Research Unit ‘Genomics to Enhance Microbiology Screening’: MXF, KK, KR, RM, OETM, SB, SS, JBr, TG, PS, and HH. The study was also supported by the Swedish Society for Medical Research (grant number PG-23-0435) and the Gothenburg Society for Medicine (grant number GLS-1001038). SFL is funded by a Wellcome Doctoral Training Fellowship (grant number 220549/Z/20/Z); this research was funded in part by the Wellcome Trust. MFP and LH acknowledge funding from Suomen Lääketieteen säätiö and Medicinska Understödsföreningen Liv & Hälsa. KH and ZD acknowledge funding from the Jane and Aatos Erkko Foundation. TG is supported by an Investigator Grant (GNT2025445) from the National Health and Medical Research Council, Australia (NHMRC). PCM receives core funding from the Francis Crick Institute (ref CC2223) and acknowledges funding support from the University College London NIHR Biomedical Research Centre (BRC). MAA is supported by a Sir Henry Dale Fellowship jointly funded by the Royal Society and Wellcome Trust (220171/Z/20/Z). The funding bodies had no role in the study’s design, data collection, analysis, or manuscript writing. For the purpose of Open Access, the author has applied a CC BY public copyright licence to any Author Accepted Manuscript version arising from this submission.

## Author contributions

Conceptualisation: HH, PS, MXF. Formal analysis: MXF, HH, PS, WLI. Investigation: MXF, MFP, SFL, JR, KK, KR, RM, OETM, LF, SB, JBo, JBS, ZD, CK, HC, MB, LH, SS, GA, KH, MAA, EN, JBr, PM, TG. Writing – Original Draft: MXF. Writing – Review & Editing: all authors. Supervision: HH, PS, MA. All authors approved the final version of the submitted manuscript.

## Data Availability Statement

The data supporting the findings of this study are included in the manuscript. Raw sequencing data have been deposited in the NCBI BioProject accession number PRJNA1364201, including BioSample accession numbers SRR36116996 to SRR36117201. Consensus full-length genome sequences have been deposited in GenBank under accession numbers PV786119 to PV786132.

