## supplementary material for "Next-Generation Sequencing Methods for Sensitive Hepatitis B Viral Genome Analysis: A European Study"

**Further details of NGS protocols.**

Details of most individual protocols have been previously published and are referenced where applicable below. The protocol for PCR-A is described below. Although the purpose of this study was not to compare the impact of specific methodological factors, a simplistic overview of the protocols is provided in Table S1. Nucleic acid extractions were performed at individual laboratories, except for three protocols for which extractions were conducted separately in the coordinating laboratory in Oxford (PCR-A, PCR-B, PCR-2).

*MTG-A*: The laboratory workflow is found at this reference^5^, where plasma samples were extracted using the QIAsymphony DSP Virus/Pathogen Mini Kit according to the manufacturer’s instructions. The bioinformatics pipeline for HBV alignment and coverage, including deduplication details, is found in this reference^6^. The bioinformatics pipeline metaMix for co-detection of viruses is found at this reference^7^.

*MTG-B/TAC-A/TAC-B*: The laboratory and bioinformatics workflows, including the bioinformatics workflow for generating the co-detection data and read deduplication, are found in this reference^8^. For read processing, the mapped fragment length (TLEN) threshold remained at the pipeline default of 40 nucleotides. Trim settings remained at pipeline defaults, including removal of Illumina adapters. The minimum fragment length to include reads was 36 nucleotides. The HBV reference set comprises 14 full-length genomes: 12 cluster-derived consensus sequences and 2 singletons, generated from 462 HBV sequences to represent the full diversity of known HBV genomes.

The TAC-A protocol, where a combined cDNA and gDNA protocol was used, was identical to the referenced cDNA-only protocol, except that 50% of the cDNA was replaced with gDNA. TAC-B utilised the cDNA-only protocol, and pre-capture libraries were sequenced for the MTG-B protocol. Thus, TAC-B and MTG-B samples underwent the same sample processing before the capture step. The cDNA protocol does not include a DNase treatment step; therefore, both gDNA and cDNA are carried forward for library preparation. The TAC-A and TAC-B protocols were designed for both RNA and DNA viruses and thus were the only protocols in this study to contain a cDNA synthesis step (which may additionally sequence replication intermediates in addition to the DNA reservoir), in contrast to TAC-C, which was designed for DNA viruses only.

*TAC-C*: The laboratory workflow is found in this reference^9^, and the bioinformatics workflow, TracesPipe, is found in this reference^10^. The minimum fragment length for raw read processing was 25 nucleotides, and deduplication was handled by samtools rmdup in TracesPipe. The TracesPipe pipeline uses FALCON, a compression-based metagenomic tool, to guide reference selection for alignment-based assembly. FALCON compares processed reads to a database of 7,555 HBV genomes and identifies the most similar genome for each sample. The database consisted of all Genbank entries labelled with a taxon ID descendant from txid10239 (viruses) as of July 11th 2019, filtered by sequence definition and length to complete HBV genomes.

*PCR-A*: The PCR protocol consists of a nested approach. The first PCR spans the genome, with four nested amplicons subsequently generated, 4-1018, 593-1667, 1451-2510 and 2416-3218, respectively (see diagram below). For the first PCR, the total PCR reaction volume is 25 μL, including 5 μL of DNA input per reaction. For the second PCR, the total PCR reaction volume is 25 μl, including 1 μl first-round amplicon per reaction.

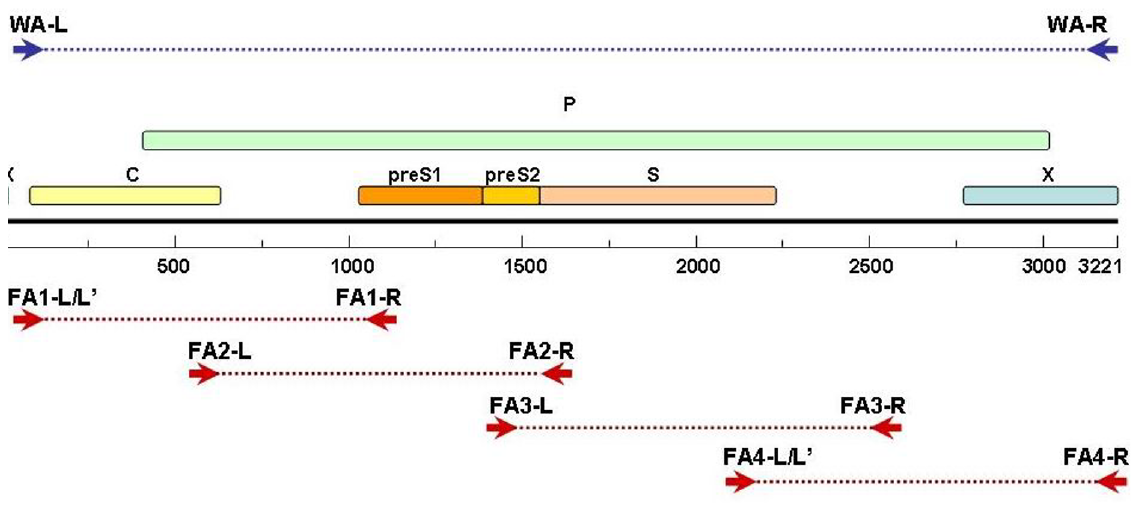

Primer sequences:

WA-L	ACTGTTCAAGCCTCCAAGCTGTGC

WA-R	AGCAAAAAGTTGCATGGTGCTGGT

FA1-L	TTTCACCTCTGCCTAATCATCTC

FA1-L'	TTT ACCTCTGCCTAATCATCTC

FA1-R	TCTTGTTCCCAAGAATATGGTG

FA2-L	GCGTCGCAGAAGATCTCAAT

FA2-R	TTGAGAGAAGTCCACCACGAG

FA3-L	CTGCTGGTGGCTCCAGTT

FA3-R	GCCTTGTAAGTTGGCGAGAA

FA4-L	GTATTGGGGGCCAAGTCTGT

FA4-L'	GTATTGGGGGCCAAATCTGT

FA4-R	AAAAAGTTGCATGGTGCTG

Equimolar pooling of amplicons from each sample was performed before library preparation with Nextera XT kit, following the manufacturer’s instructions. 1 ng (5 μl of 0.2 ng/μl) pooled amplicon input was used for each library preparation reaction.

For initial bioinformatics analysis, a modified version of the ADPU (UCL Hospitals) HBV Bioinformatics Pipeline was utilised^11^, incorporating deduplication but excluding the variant analysis step. This pipeline is designed for the assembly and analysis of HBV genomes from paired-end Illumina sequencing data:

1. Raw Read Processing (Tool: Trimmomatic v0.39)

Settings:

- Cores: 8
- Mode: Paired-end
- Adapter removal: ILLUMINACLIP:<adapter_file>:2:10:7
- Quality trimming: LEADING:10; TRAILING:10; SLIDINGWINDOW:4:30; MINLEN:50 [reads shorter than 50 nucleotides are removed]

2. Host Decontamination (Tool: SMALT v0.7.6)

- Reads are aligned to a decoy reference consisting of the human genome and HBV reference sequences
- Paired-end mapping with -x -y 0.5 -i 500 -n <cores>
- Only non-human read pairs are retained for downstream processing

3. Decontaminated Read Extraction (Tool: samtools)

- Extract properly paired reads from SAM file into two FASTQ files using samtools view -bhf and samtools bam2fq
- Remove PCR duplicates and redundant reads using samtools markdup

4. De Novo Assembly (Tool: IVA (Iterative Virus Assembler) [Docker: sangerpathogens/iva])

Settings:

- --max_contigs 50
- Uses filtered paired-end reads
- Uses the same adapter and primer files as preprocessing
- Runs in a Docker container with volumes mounted for data and scripts

5. Contig Alignment and Best Reference Selection (Tool: LASTZ)

- Contigs aligned to a reference scaffold database using LASTZ with --ambiguous=IUPAC
- The database contains 958 full-length HBV genomes: the Refseq sequences, other references for subtypes, and the rest from Genbank, filtered by sequence length and date submitted (approximately 2020-2024; all sequences were manually inspected).
- Best matching reference genome is selected using lastz_bestref.NO_BLAST.pl

6. Contig-to-Reference Alignment Analysis (Tool: lastz_analyser.WITH_REVCOMP.pl)

- Determines the relationship between contigs and the best reference
- Cutoff: 50,000 bp
- Allows reverse complement alignment

7. Initial Consensus Generation (Tool: genome_maker2b.pl)

- Combines mapped reads and contigs to generate a draft genome
- Aligns reads to contigs using SMALT
- Generates a BAM file and mpileup for consensus building

8. Consensus Polishing (First Round) (Tool: cons_mv.pl)

Settings:

- Frequency cutoff: 0.01
- Minimum depth: 100 (overall), 20 (variant), 80 (consensus)
- Sliding window: 300 bp

(Tool: N_remover_from_consensus.pl)

- Removes Ns from consensus with a cutoff of 46

9. Consensus Polishing (Second Round)

- Same steps as above are repeated on the first polished consensus to yield a final polished consensus (consensus2)

*PCR-B/PCR-2:* The laboratory and bioinformatics workflows can be found at this reference^12^.

*PCR-1*: The laboratory workflow can be found at this reference^13^, and the bioinformatics workflow can be found at this reference^14^, where Guppy v4.2.0 was used for base-calling. The deduplicate tool in samtools was used, but none of the reads for this study were duplicates, in agreement with the non-cluster-based sequencing performed in Nanopore.

**Co-detection of other virus species by PCR.** Multiplex real-time PCR was performed to detect all human herpesviruses (HHV) as previously described^1^. Real-time PCR for human polyomavirus 6 (HPyV6), JC polyomavirus (JCPyV), and Merkel cell polyomavirus (MCPyV) was performed as previously described^2^.

For anellovirus, a previously described nested PCR protocol to amplify alphatorquevirus, betatorquevirus, and gammatorquevirus separately ^3^ was modified by using GoTaq G2 DNA Polymerase (Promega) in a total 25 μl reaction volume with 5 μl of extracted DNA (extraction protocol using 5 ml of plasma^4^) in the first-round reaction and 2 μl of first-round template in the second-round reaction. Further, the annealing temperature was modified to 58^o^C and the number of cycles during second-round amplification was altered to 35. Samples positive for the specific torqueviruses by nested PCR underwent confirmatory secondary PCR using SYBR Select Master Mix (Thermofisher Scientific). Reaction volumes were 20 μL, with 2 μL of first-round template with 200 nM primers. Cycle conditions were 95^o^C 5 min, 40 cycles of 95°C 15 sec, 58°C 1 min, and a dissociation curve. A random selection of amplicons was also sent for Sanger sequencing as previously described^4^.

**Table S1**. Protocol details of the NGS protocols analysed, including the sequencing workflows and analysis pipelines (until the point of genome assembly) for each method. Effective test volume was calculated as [library prep input volume]/[extraction output volume]*[extraction input volume], where TAC-A, TAC-B and MTG-B also accounted for reductions in effective test volume through cDNA synthesis, and PCR volumes were also accounted for in PCR-based protocols. Numerical superscripts next to method identifiers in the column headings indicate where the same laboratory performed the differing protocols.

|  | **MTG-A** | **MTG-B^1^** | **TAC-A^1^** | **TAC-B^1^** | **TAC-C** | **PCR-A** | **PCR-B^2^** | **PCR-1** | **PCR-2^2^** |
| --- | --- | --- | --- | --- | --- | --- | --- | --- | --- |
| **Technology** | Illumina | Illumina | Illumina | Illumina | Illumina | Illumina | Illumina | ONT | ONT |
| **Pre-extraction concentration** | No | PEG Virus Precipitation | No | PEG Virus Precipitation | No | No | No | No | No |
| **Nucleic acid extraction** | QIA-symphony DSP Virus/Pathogen Mini Kit | Roche Large Volume Kit | Roche Large Volume Kit | Roche Large Volume Kit | QIAamp DNA Mini Kit | Roche Large Volume Kit* | Roche Large Volume Kit* | Roche Large Volume Kit | Roche Large Volume Kit* |
| **Input/output volume** | 200 μL /  60 μL | 20,000 μL / 50 μL | 5,000 μL / 50 μL | 20,000 μL / 50 μL | 400 μL /  60 μL | 5,000 μL /  50 μL | 5,000 μL /  50 μL | 5,000 μL /  50 μL | 5,000 μL /  50 μL |
| **Pre-prep PCR amplification (volume of extract added, total reaction volume)** | No | No | No | No | No | Yes  Round 1: 5 μL, 25 μL  Round 2 (x4): 1 μL, 25 μL | Yes  2x (7 μL, 25 μL), followed by clean-up into 20 μL | Yes  2x (5 μL, 25 μL) | Yes,  2x (7 μL, 25 μL), followed by clean-up into 20 μL |
| **Library prep input volume** | 26 μL | 5 μL | 5 μL | 5 μL | 50 μL | 5 μL  (0.2 ng/μL) | 8.3 μL | 10 μL | 8.3 μL |
| **Effective test volume** | 87 μL | 800 μL | 350 μL | 800 μL | 333 μL | ~4 μL | 581 μL | 200 μL | 581 μL |
| **rRNA depletion** | No | No | No | No | No | No | No | No | No |
| **Human DNA depletion** | No | No | No | No | No | No | No | No | No |
| **cDNA synthesis** | No | Yes | Yes | Yes | No | No | No | No | No |
| **Library prep kit** | NEBNext Ultra II FS (DNA) | Twist (cDNA) | Twist (half DNA & half cDNA) | Twist (cDNA) | Roche Hyperprep (DNA) | Nextera XT (DNA) | NEBNext Ultra II FS  (DNA) | SQK-RBK114.96 (DNA) | SQK-NBD114.96(DNA) |
| **Probe capture** | No | No | Yes | Yes | Yes | No | No | No | No |
| **Random amplification cycles** | N/A | 16 | 16 | 16 | 30 (2x 15) | 12 | 0 | 30 | 0 |
| **Average gigabases (Gb) or megabases (Mb) sequenced per sample**  **Platform** | 5.03 Gb    NextSeq 2000 | 208.2 Mb  NovaSeq X | 49.6 Mb  NovaSeq X | 105.1 Mb  NovaSeq X | 2.5 Gb  NovaSeq X | 317.3 Mb  MiSeq V2 | 47.4 Mb  NovaSeq X Plus | 2.2 Mb  MinION | 312.7 Mb  MinION |
| **Sequencing strategy** | Paired end 150 | Paired end 150 | Paired end 150 | Paired end 150 | Paired end 150 | Paired end 250 | Paired end  150 | N50:  1.06 kb | N50:  600 bp |
| **Read filtering** | nf-core taxprofiler (MetaMix was used for detecting additional viruses) | Kraken2, FastQC, Trimmomatic, BWA-mem2, custom script for aggregating reads by reference organism, samtools, viralconsensus | Castanet | Castanet | TracesPipe | FASTQC, Trimmomatic, Smalt | QUASR, CutAdapt, Skewer, Bowtie2 | See supplementary material. | dna_r10.4.1_e8.2_400bps_hac@v5.0.0_SQK-NBD114-96, hbv-fieldbioinformatics |
| **Assembly method** | Reference-based mapping | Reference-based mapping | Reference-based mapping | Reference-based mapping | Bowtie 2 and metaSPAdes | IVA, de novo using custom scripts | Reference guided to closest reference | Minimap2 | Reference guided to closest reference |
| **Reference genomes** | All complete RefSeq genomes and NC_003977.2 | 14 full-length cluster-derived consensus sequences | 14 full-length cluster-derived consensus sequences | 14 full-length cluster-derived consensus sequences | All filtered Genbank entries | Database with 958 genomes of different subtypes | 44 references covering all known HBV subgenotypes^32^ | Reference genome for each genotype A to I | 44 references covering all known HBV subgenotypes^32^ |

*These extractions were performed separately in the central coordinating centre, and eluates (or spin columns before the elution step) were sent to the laboratories instead of plasma.

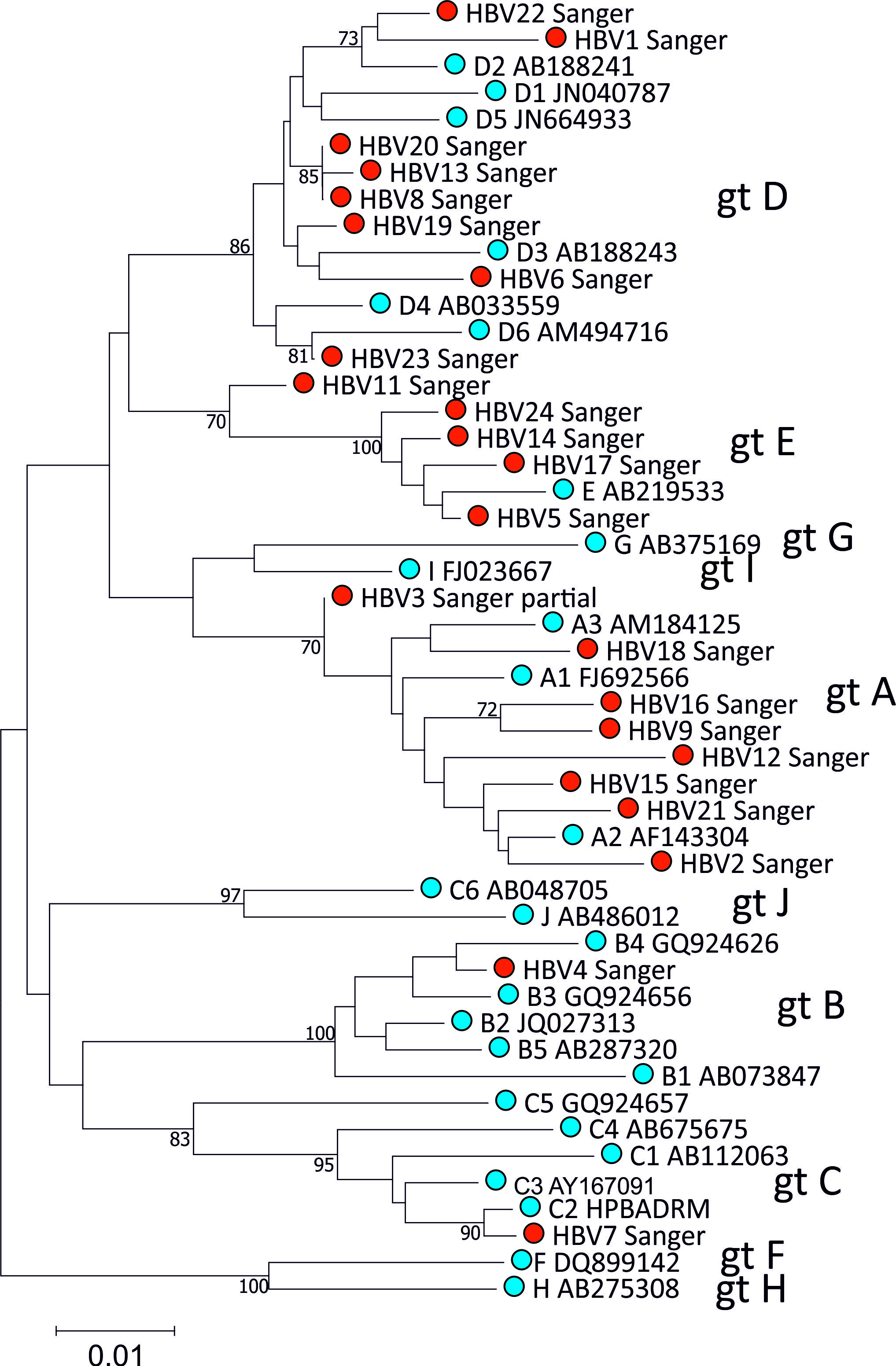

**Figure S1.** Phylogenetic tree of the sample panel (coloured in blue), with reference sequences of HBV subgenotypes (coloured in red). Gt = genotype. Evolutionary history was inferred using the Neighbor-Joining method with 100 bootstrap replicates. Evolutionary distances were computed using the Maximum Composite Likelihood method, conducted in MEGA7.

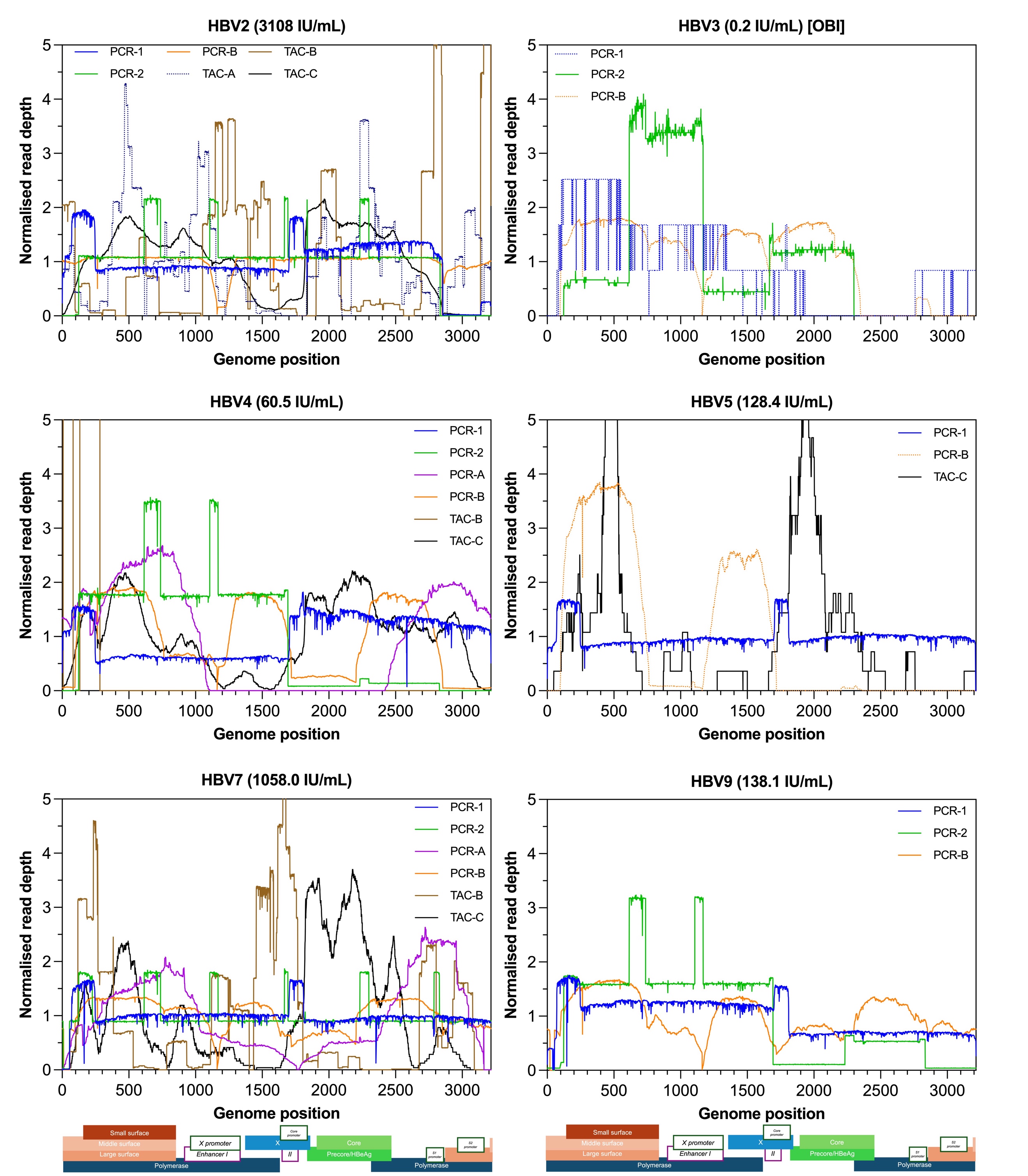

**Figure S2**. Coverage of the HBV genome for each sample and sequencing method. Only sequences with >1000 total HBV-specific bases read and >50% genome coverage were included. Genome positions were based on the D00330 reference sequence. A genome diagram of HBV drawn to the x-axis scale shows the gene positions in shaded boxes and regulatory regions in unshaded boxes. Viral loads are shown beside the sample name and occult HBV infection (OBI) status.

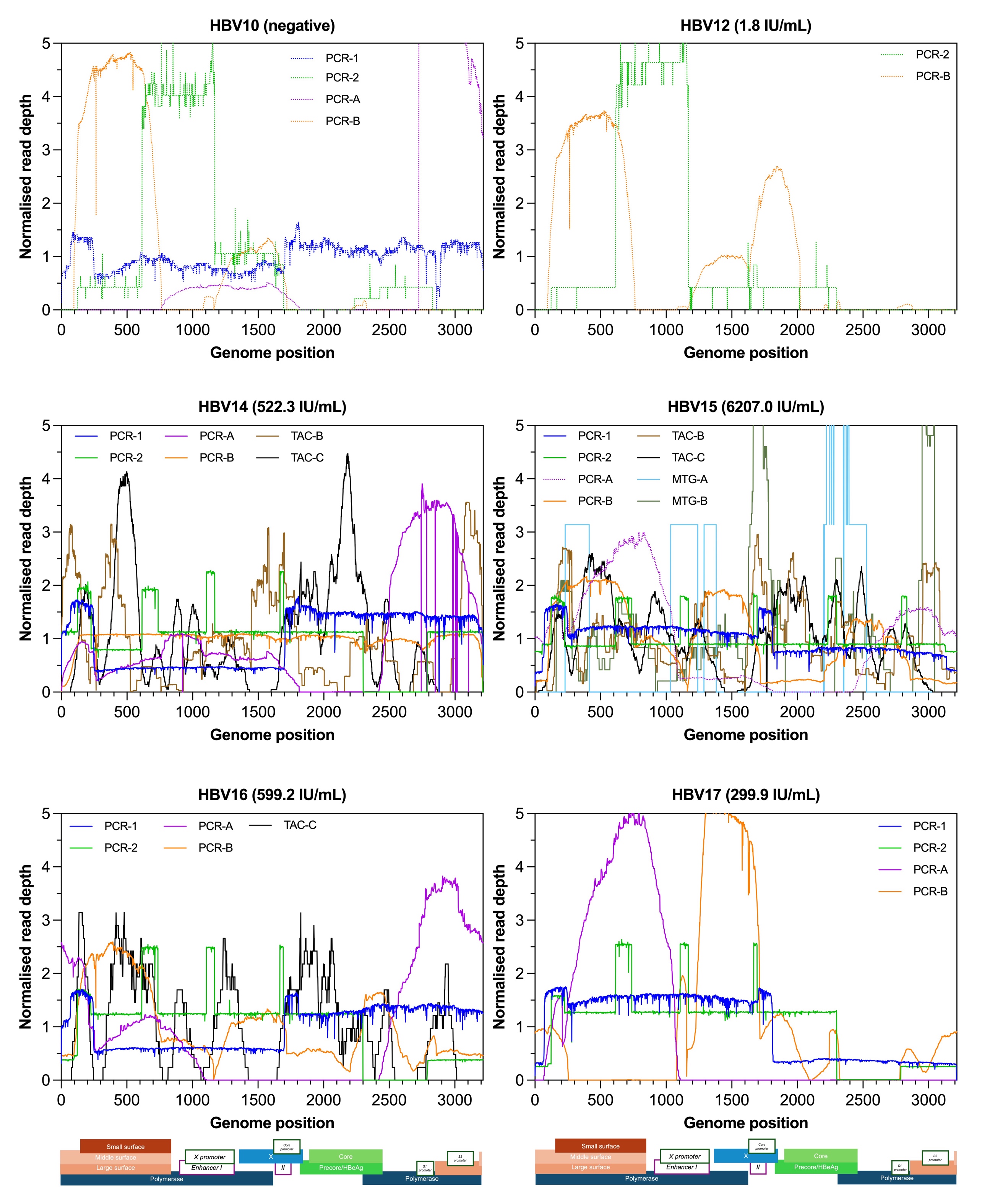

**Figure S2** (continued).

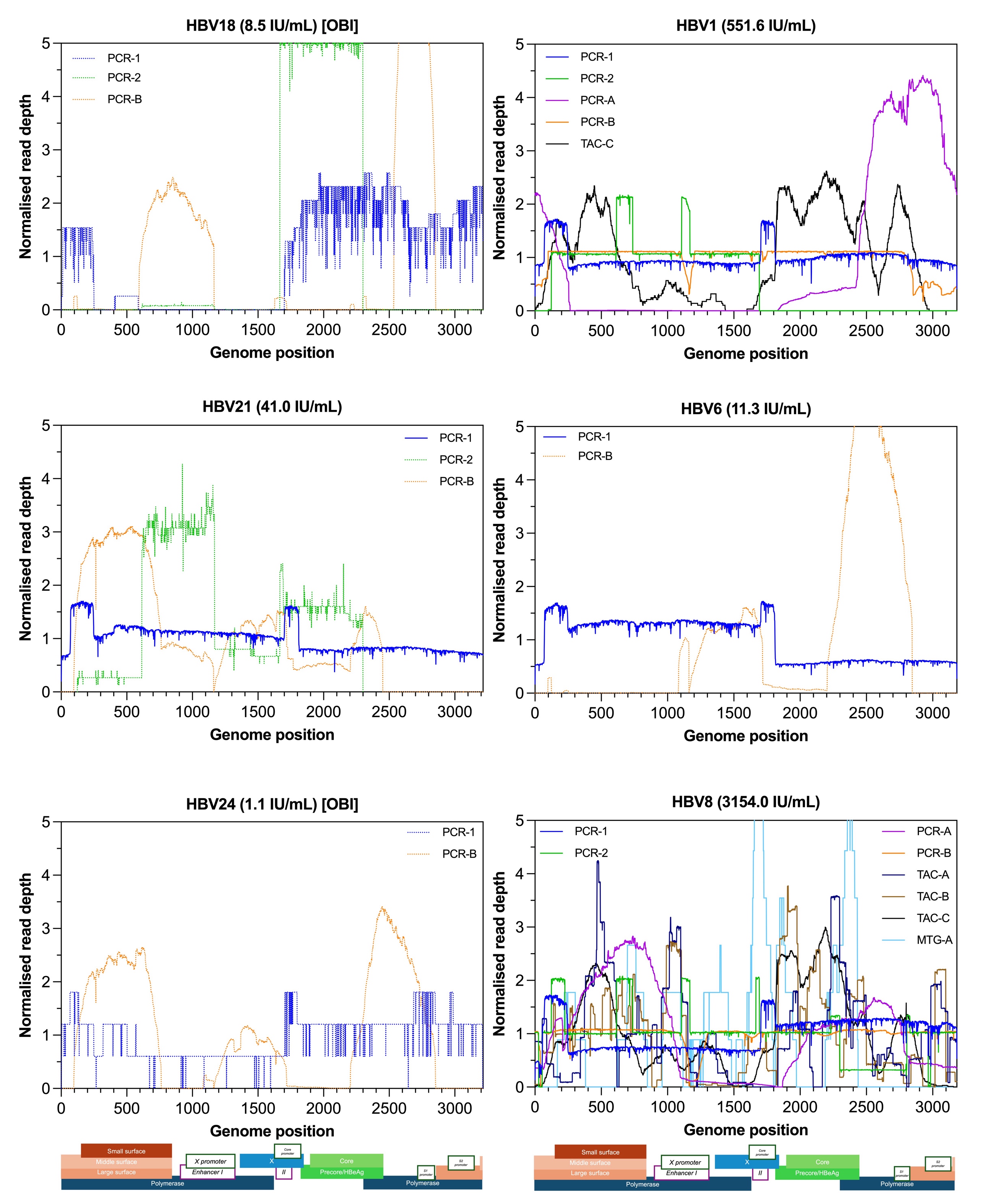

**Figure S2** (continued).

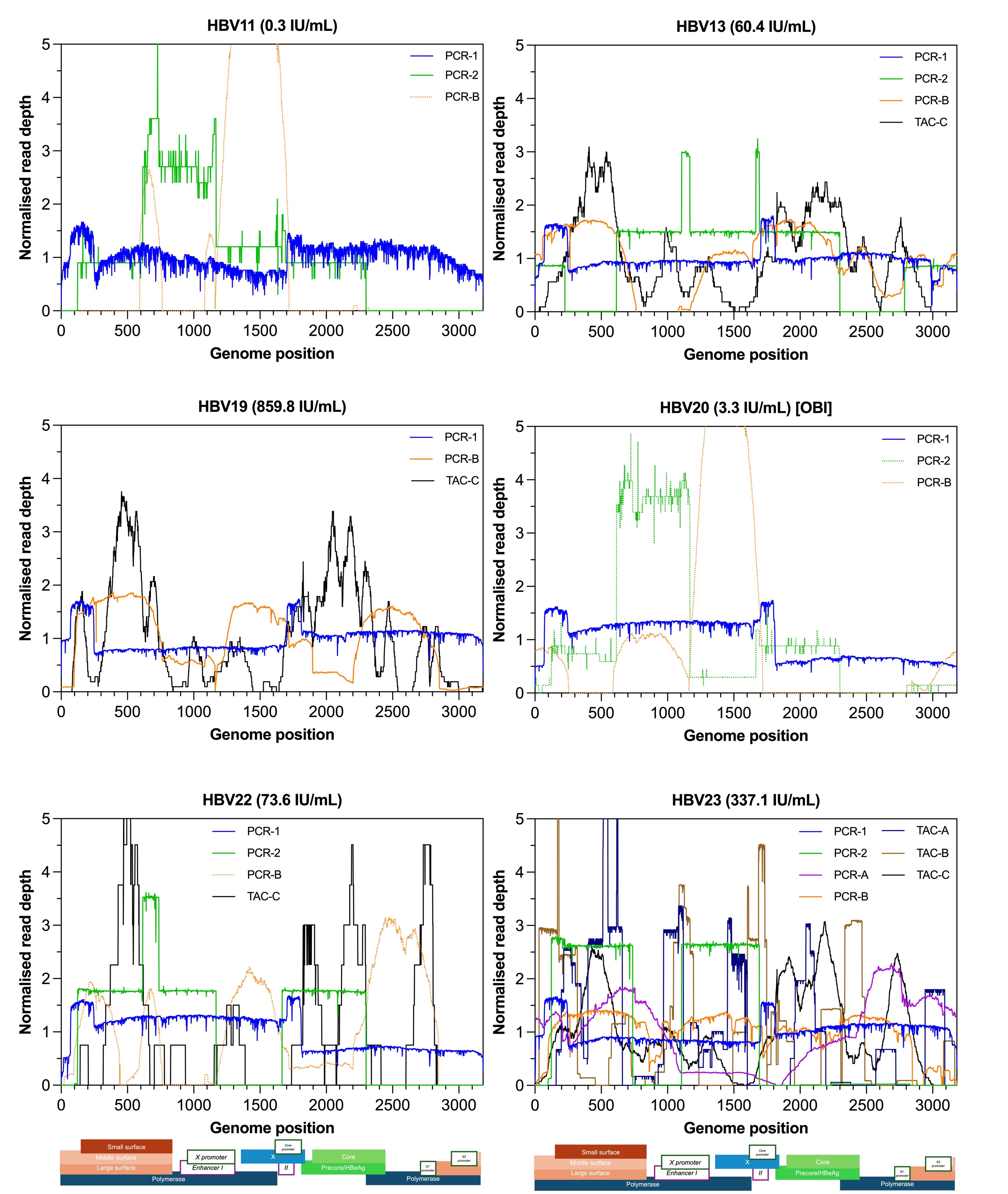

**Figure S2** (continued).

**Table S2**. Details of primers used for sequencing of HBV amplicons, indicating the start and end nucleotide positions of the amplified regions. Outer primer sets are provided in the above cell, while inner primers are indicated in the cell below. Nucleotide positions are numbered according to the D00330.1 reference sequence.

| **Region** | **Primer sequence (5’ to 3’)** | **Nucleotide positions of amplified region** | **Length of region** |
| --- | --- | --- | --- |
| S | CATCAGGAYTCCTAGGACCCCT; GAGGCATAGCAGCAGGATGMAGAGG;  GACTTCTCTCAATTTTCTAGGGG;  AGTAAACTGAGCCAAGAGAAACGG | 219-375 &  400-637 | 395  (157 & 238) |
|  | CGTGTTACAGGCGGKGTKTTTCTTGT;  ATGATAAAACGCCGCAGACACATC;  GATGTGTCTGCGGCGTTTTATCAT;  ACGGACTGAGGCCCACTCCCATAG |  |  |
| SP | ATCCMGAYTGGGACYTCAA;  CGTTGCCKDGCAACSGGGTAAAGG | 3212-969 | 973 |
|  | TCATCCTCAGGCCATGCAGT;  GACACACTTTCCAATCAATNGG |  |  |
| XC | TCTTGCCCA AGGTCTTACAT;  TCCCACCTTATGAGTCCAAG | 1673-2370 | 698 |
|  | ATAAGMGGACTMTTGGACT;  CAGCGAGGCGAGGGAGTTCTTCTT |  |  |

**Table S3.** Overview of the detection of HBV genotypes in each protocol for each sample. Each genotype (A to E) is shaded in a different colour. The absence of sequences was shaded in black. “-“ indicates where sequence length was insufficient to determine HBV genotype. Samples are ranked from highest to lowest geometric mean viral load. “*” indicates where sequences were not comparable for HBV18 because of limited overlap.

| **Genotype** |  |  |  |  |  |  |
| --- | --- | --- | --- | --- | --- | --- |
|  | No sequence | A | B | C | D | E |

|  |  | **PCR (Nanopore)** | | **PCR (Illumina)** | | **Target capture** | | | **Metagenomics** | |  |  |
| --- | --- | --- | --- | --- | --- | --- | --- | --- | --- | --- | --- | --- |
| **Sample** | **Viral load (IU/mL)** | **PCR-1** | **PCR-2** | **PCR-A** | **PCR-B** | **TAC-A** | **TAC-B** | **TAC-C** | **MTG-A** | **MTG-B** | **Within-sample**  **consensus** | **Genotype**  **assigned** |
| **HBV15** | **6207.0** | A | A | E | A | A | A | A | A | A | **Y** | **A2** |
| **HBV8** | **3154.0** | D | D | D | D | D^b^ | D | D | D | D | **Y** | **D1** |
| **HBV2** | **3108.0** | A | A | NS | A | D^b^ | A | A | NS | A | **Y** | **A2** |
| **HBV7** | **1058.0** | C | C | C | C | C | C | C | C | C | **Y** | **C1** |
| **HBV19** | **859.8** | D | D | NS | D | D | D | D | D | NS | **Y** | **D3** |
| **HBV16** | **599.2** | A | A | A | A | A | NS | A | NS | NS | **Y** | **A1** |
| **HBV1** | **551.6** | D | D | D | D | D | D | D | - | NS | **Y** | **D2** |
| **HBV14** | **522.3** | E | E | E | E | E | E | E | NS | E | **Y** | **E** |
| **HBV23** | **337.1** | D | D | D | D | D | D | D | NS | D | **Y** | **D4** |
| **HBV17** | **299.9** | E | E | E | E | NS | E | E | NS | NS | **Y** | **E** |
| **HBV9** | **138.1** | A | A | A | A | A | A | A | A | NS | **Y** | **A1** |
| **HBV5** | **128.4** | E |  | NS | C | NS | NS | E | NS | NS | **N** | **E** |
| **HBV22** | **73.6** | D | D | NS | C | NS | NS | D | NS | NS | **Y** | **D2** |
| **HBV4** | **60.5** | B | B | B | B | B | B | B | B | NS | **Y** | **B4** |
| **HBV13** | **60.4** | D | D | NS | D | NS | NS | D | D | NS | **Y** | **D1** |
| **HBV21** | **41.0** | A | D | NS | C^a^ | NS | A | A | NS | NS | **N** | **A** |
| **HBV6** | **11.3** | D |  | NS | B | NS | D | D | NS | D | **N** | **D3** |
| **HBV18** | **8.5** |  | A* | NS | B* | NS | NS | NS | NS | NS | **N** | **A** |
| **HBV20** | **3.3** | D | A | NS | B | NS | NS | D | - | NS | **N** | **D** |
| **HBV12** | **1.8** |  | E | NS | B | NS | NS | D | NS | NS | **N** | **A** |
| **HBV24** | **1.1** |  |  | NS | C^a^ | NS | NS | NS | NS | NS | **N** | **E** |
| **HBV11** | **0.3** | D | D | NS | C^a^ | NS | NS | NS | NS | NS | **N** | **D** |
| **HBV3** | **0.2** |  | E | NS | C | NS | NS | E | NS | NS | **N** | **A3** |
| **Negative**  **(HBV10)** | **-** | D | A | D | B | NS | NS | NS | NS | NS | **N** | **-** |

**Table S4**. Overview of the detection of HBV in sample sets, when majority consensus bases were called at a threshold of 1 base per nucleotide site for all methods. The percentage genome completeness is given in each cell. The left-hand side of the table with blue headers shows the sequences obtained from the NGS methods, whilst the right-hand side (calamine colour) shows the sequences obtained by Sanger sequencing (length of S amplicon = 395 nucleotides; SP = 973 nucleotides; XC = 698 nucleotides). Assembled sequences were assembled for each sample from the individual sequences of the NGS methods that had around 100% genome coverage and were genetically similar. Assembled sequences were assigned to HBV genotypes. The percentage genetic relatedness of the sequences from each method was compared to the within-sample consensus sequence, where assembled sequences similar to the within-sample consensus were shaded on a grayscale, and different strains (differing in >5% nucleotide sequence from the within-sample consensus) were shaded in a pink scale. The absence of sequences was shaded in black. Where within-sample consensus sequences could not be assembled (i.e. no two complete sequences were similar), genotypes were defined from Sanger sequences, and the NGS sequences were instead compared to one another. Samples are ranked from highest to lowest geometric mean viral load. For TAC-B: library preparation failed for sample HBV13; samples HBV4, HBV9, and HBV19 had reads of poor quality for the original pipeline to process, so the mean base quality was dropped below 1.0. #: For PCR-1, a negative water control was tested instead of the negative plasma control. Alphabetical superscripts (a, b, c) represent identical sequences. Numerical superscripts next to method identifiers indicate where the same laboratory performed the protocols.

|  |  | **PCR (Nanopore)** | | **PCR (Illumina)** | | **Target capture** | | | **Metagenomics** | |  |  | **Sanger sequencing** | | |
| --- | --- | --- | --- | --- | --- | --- | --- | --- | --- | --- | --- | --- | --- | --- | --- |
| **Sample** | **Viral load (IU/mL)** | **PCR-1** | **PCR-2^1^** | **PCR-A** | **PCR-B^1^** | **TAC-A^2^** | **TAC-B^2^** | **TAC-C** | **MTG-A** | **MTG-B^2^** | **Within-sample**  **consensus** | **Genotype**  **assigned** | **S** | **SP** | **XC** |
| **HBV15** | **6207.0** | 100% | 100% | 91% | 100% | 11% | 100% | 90% | 25% | 72% | **Y** | **A2** | 12% | 30% | 22% |
| **HBV8** | **3154.0** | 100% | 100% | 98% | 100% | 91%^c^ | 97% | 99% | 50% | 14% | **Y** | **D1** | 12% |  |  |
| **HBV2** | **3108.0** | 100% | 91% | NS | 100% | 90%^c^ | 81% | 100% | NS | 25% | **Y** | **A2** | 12% |  | 22% |
| **HBV7** | **1058.0** | 100% | 100% | 98% | 99% | 17% | 83% | 93% | 11% | 25% | **Y** | **C1** | 12% | 29% | 22% |
| **HBV19** | **859.8** | 100% | 49% | NS | 100% | 22% | 4% | 88% | 10% | NS | **Y** | **D3** | 12% | 31% | 22% |
| **HBV16** | **599.2** | 100% | 85% | 59% | 100% | 9% | NS | 75% | NS | NS | **Y** | **A1** | 12% |  |  |
| **HBV1** | **551.6** | 100% | 86% | 76% | 100% | 6% | 38% | 88% | 4% | NS | **Y** | **D2** | 12% | NS | NS |
| **HBV14** | **522.3** | 100% | 100% | 92% | 100% | 9% | 95% | 80% | NS | 49% | **Y** | **E** | 12% | 30% |  |
| **HBV23** | **337.1** | 100% | 86% | 99% | 100% | 64% | 71% | 93% | NS | 16% | **Y** | **D4** | 12% | 31% | 22% |
| **HBV17** | **299.9** | 100% | 100% | 96% | 92% | NS | 14% | 50% | NS | NS | **Y** | **E** | 12% | 30% |  |
| **HBV9** | **138.1** | 100% | 100% | 33% | 99% | 9% | 8% | 35% | 2% | NS | **Y** | **A1** | 12% | 30% |  |
| **HBV5** | **128.4** | 100% | 35% | NS | 89% | NS | NS | 66% | NS | NS | **N** | **E** | 12% | 30% |  |
| **HBV22** | **73.6** | 100% | 83% | NS | 86% | NS | NS | 54% | NS | NS | **Y** | **D2** | 12% | 31% |  |
| **HBV4** | **60.5** | 100% | 85% | 59% | 100% | 35% | 7% | 100% | 12% | NS | **Y** | **B4** | 12% | 30% | 22% |
| **HBV13** | **60.4** | 100% | 85% | NS | 98% | NS | NS | 91% | 5% | NS | **Y** | **D1** | 12% | 31% | 22% |
| **HBV21** | **41.0** | 100% | 68%^a^ | NS | 74% | NS | 7% | 43% | NS | NS | **N** | **A** | 12% | 30% |  |
| **HBV6** | **11.3** | 100% | 33% | NS | 90% | NS | 3% | 28% | NS | 3% | **N** | **D3** | 12% | 31% |  |
| **HBV18** | **8.5** | 60% | 67% | NS | 88% | NS | NS | NS | NS | NS | **N** | **A** | 12% |  |  |
| **HBV20** | **3.3** | 100% | 82%^a^ | NS | 81% | NS | NS | 26% | <1% | NS | **N** | **D** | 12% |  |  |
| **HBV12** | **1.8** | 15% | 67% | NS | 97% | NS | NS | 25% | NS | NS | **N** | **A** | 12% |  |  |
| **HBV24** | **1.1** | 99% | 33%^b^ | NS | 88% | NS | NS | NS | NS | NS | **N** | **E** | 12% |  |  |
| **HBV11** | **0.3** | 100% | 68% | NS | 85% | NS | NS | NS | NS | NS | **N** | **D** | 12% |  |  |
| **HBV3** | **0.2** | 71% | 68% | NS | 91% | NS | NS | 5% | NS | NS | **N** | **A3** | 5% |  |  |
| **Negative**  **(HBV10)** | **-** | 100%^#^ | 67%^b^ | 48% | 88% | NS | NS | NS | NS | NS | **N** | **-** | NS | NS | NS |

| **Within-sample consensus** | **Shading key** | | | | | |
| --- | --- | --- | --- | --- | --- | --- |
| **Y (% sequence identity to within-sample consensus)** |  |  |  |  |  |  |
|  | No sequence | ≥85% | ≥90% | ≥95% | ≥98% | ≥99.5% |
| **N (similarity of sequences)** |  |  |  |  |  |  |
|  | No sequence | Different sequence to others | Similar sequence |  |  |  |

**Table S5.** Specific criteria used by protocols to call majority bases and positive detection, in addition to the read criteria already detailed in the supplementary information.

| **Protocol** | **Minimum depth for majority base calls** | **Other criteria for positive detection** |
| --- | --- | --- |
| MTG-A | 1 | - |
| MTG-B, TAC-A, TAC-B | 3 | Minimum genome coverage 10% |
| TAC-C | 1 | - |
| PCR-A | 80 | - |
| PCR-B | 5 | - |
| PCR-1 | 10 | Minimum amplicon threshold of 100 for resistance mutation assessment in the RT or for genotyping  10 times more reads in the sample than in the negative control |
| PCR-2 | 20 | - |
